# Polygenic score informed by genome-wide association studies of multiple ancestries and related traits improves risk prediction for coronary artery disease

**DOI:** 10.1101/2023.03.03.23286649

**Authors:** Aniruddh P. Patel, Minxian Wang, Yunfeng Ruan, Satoshi Koyama, Shoa L. Clarke, Xiong Yang, Catherine Tcheandjieu, Saaket Agrawal, Akl C. Fahed, Patrick T. Ellinor, Genes & Health Research Team, the Million Veteran Program, Phillip S. Tsao, Yan V. Sun, Kelly Cho, Peter W. F. Wilson, Themistocles L. Assimes, David A. van Heel, Adam S. Butterworth, Krishna G. Aragam, Pradeep Natarajan, Amit V. Khera

## Abstract

Accurate stratification of coronary artery disease (CAD) risk remains a critical need. A new polygenic score (GPS_Mult_) incorporates CAD genome-wide association data across five ancestries (>269,000 cases, >1,178,000 controls) with genetic association data for ten CAD risk factors. GPS_Mult_ associates with an OR/SD 2.14, (95%CI:2.10-2.19,P<0.001) for prevalent CAD and HR/SD 1.73 (95%CI 1.70-1.76,P<0.001) for incident CAD. When compared with the previously published GPS_2018_ in external datasets, GPS_Mult_ demonstrated 73%, 46%, and 113% increase in effect size for individuals of African, European, and South Asian ancestry, respectively, and significantly outperformed recently published CAD polygenic scores. GPS_Mult_ identifies individuals with CAD risk extremes, including the top 3% of the population at equivalent risk for a new CAD event as those with prior CAD having a second event. Integrating GPS_Mult_ with the Pooled Cohort Equations results in 7.0% [95%CI:5.9%-8.2%,P<0.001] net reclassification improvement at the 7.5% threshold. Large-scale integration genetic association data for CAD and related traits from diverse populations meaningfully improves polygenic risk prediction.

## INTRODUCTION

Coronary artery disease (CAD) remains the leading cause of death worldwide and identification of at-risk individuals remains a critical public health need.^1^ If identified early, at-risk individuals can benefit from more efficiently targeted lifestyle interventions and cholesterol-lowering medications toward lifelong risk mitigation.^2^ However, commonly used clinical risk estimators for CAD were optimized for use in middle-aged adult populations in historical cohort studies and consequently underperform in younger populations or individuals of non-European ancestries.^3–6^ As CAD is a heritable disease, leveraging the increasing amount of widely available genetic data offers additional opportunities to significantly enhance CAD risk prediction across all groups early in life, particularly at the extremes of the risk distribution.^7,8^

Polygenic scores integrate data from genome-wide association studies into a single quantitative and predictive metric of inherited risk.^9–12^. Several studies to date have stratified individuals into substantial gradients of CAD risk based on their polygenic score beyond their clinical risk factor profiles.^13–17^ Given this potential, polygenic scores are now already being deployed clinically across some biobanks and returned through direct-to-consumer testing platforms.^18,19^ The past decade has seen numerous advances in score development, however, there still remains room for improvement in their performance, particularly among individuals of non-European ancestry.^20^ Simulation studies suggest that larger sample sizes of GWAS have the potential to more accurately estimate the effect size associated with each SNP to improve scores for CAD.^21^ Polygenic scores integrating GWAS data from individuals of diverse ancestries in addition to that of the target population show relative improvement in predictive accuracy compared with methods only utilizing GWAS data from a single ancestry source.^22–26^ Furthermore, the principles of genetic correlation suggest benefit in incorporating information from GWAS of related traits to refine polygenic prediction in the trait of interest.^27–34^

Alongside considerable enthusiasm for polygenic scores to enable a new era of preventive clinical medicine is recognition of several key unmet needs before polygenic scores can be more widely implemented. First, polygenic scores have reduced predictive performance in individuals of non-European ancestry.^35^ This largely stems from relative underrepresentation of other ancestries in prior GWAS discovery cohorts. Recent efforts have focused on conducting GWAS in larger and more ancestrally diverse populations and designing methods leveraging ancestry-specific linkage disequilibrium patterns to help improve score performance.^25,36–42^ Second, although available scores associate strongly with prevalent disease, they perform less well in predicting incident disease, which would offer more clinical utility.^14^ Finally, most risk prediction models to date are based either on genetic or clinical risk factors, but better integration of these modalities and estimation of a clinically actionable risk estimate is needed.^43–45^

To address these needs, we used information from five-fold larger and more ancestrally diverse GWAS compilation compared to prior efforts along with methods leveraging commonalities in mechanistic pathways to develop a new polygenic risk score for CAD.

## RESULTS

Summary statistics from GWAS for CAD, other atherosclerotic diseases, and their risk factors across over 1.2 million individuals from multi-ancestry cohorts were aggregated to design polygenic risk scores for CAD (Figure 1, Supplementary Table 1). These scores were trained within the UK Biobank cohort in 116,649 individuals of European ancestry and then validated in the remaining independent study population of 325,991 individuals (54.3% female, 7281 African, 1,464 East Asian, 308,264 European, and 8,982 South Asian ancestry) (Supplementary Table 2).^46^ The participants in the training and validation cohorts are independent from the individuals analyzed in the previously conducted GWAS from which summary statistics were obtained. A total of 58 ancestry- and trait-specific scores were included in the GPS training analysis, with 32 scores significantly contributing to overall prediction after optimization of score selection and weighting through logistic regression (Figure 2A and 2B).

**Figure 1:**
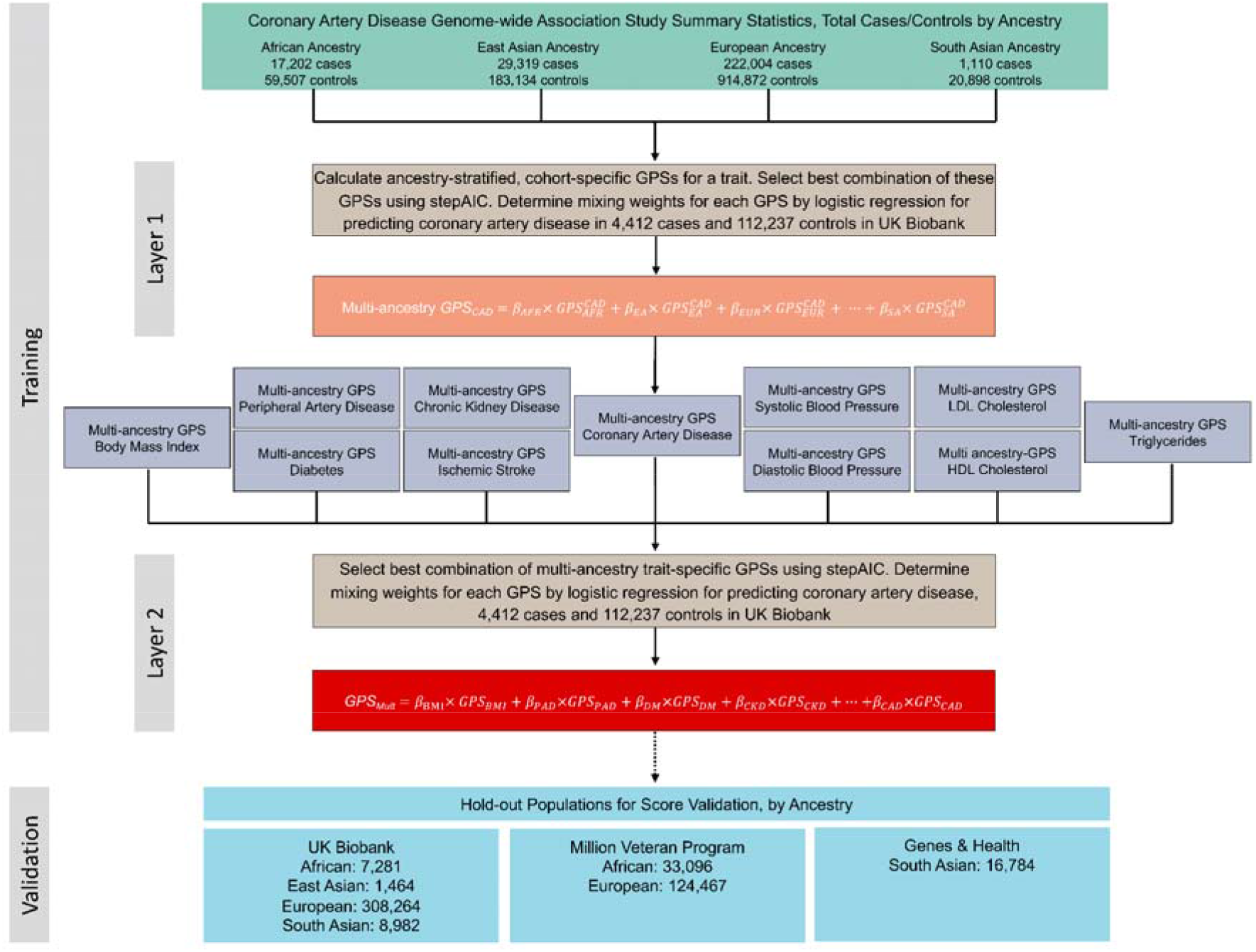
Overview of GPS_Mult_ development Polygenic scores for coronary artery disease (CAD) were constructed using ancestry-stratified, cohort-specific summary statistics from CAD and CAD-related traits, resulting in 58 GPS across all traits and ancestries. For each source trait (e.g., CAD) the best performing combination of ancestry-stratified, cohort-specific GPS was determined based on ability to predict CAD, selected using stepAIC, and their optimal mixing weights () determined using logistic regression in 116,649 individuals of European ancestry in the UK Biobank training dataset. The selected GPSs were linearly combined using these mixing weights to yield multi-ancestry scores predicting CAD from each source trait (layer 1). The best performing combination of multi-ancestry, trait-specific GPSs in predicting CAD was determined using stepAIC, and their optimal mixing weights () were determined using logistic regression in 116,649 individuals of European ancestry in the UK Biobank training dataset. The selected GPSs were linearly combined using these mixing weights to yield GPS_Mult_ (layer 2). GPS_Mult_ was validated with prediction of CAD in the UK Biobank and externally validated in Million Veteran Program and Genes & Health Studies in hold-out populations not included in score training. Ancestries: AFR – African; EA – East Asian; EUR – European; SA – South Asian. Source GWAS traits: CAD^9,39,53,57,110^, body mass index (BMI)^57,93^, ischemic stroke^57,90,111^, diabetes mellitus (DM)^91,111,112^, peripheral artery disease (PAD)^57,61,110^, chronic kidney disease (CKD)^57,63^, systolic blood pressure (SBP)^57,113^, diastolic blood pressure (DBP)^57,113^, low-density lipoprotein cholesterol (LDL)^57,62,79^, high-density lipoprotein cholesterol (HDL)^57,62,79^, triglycerides (TG)^57,62,79^.

**Figure 2:**
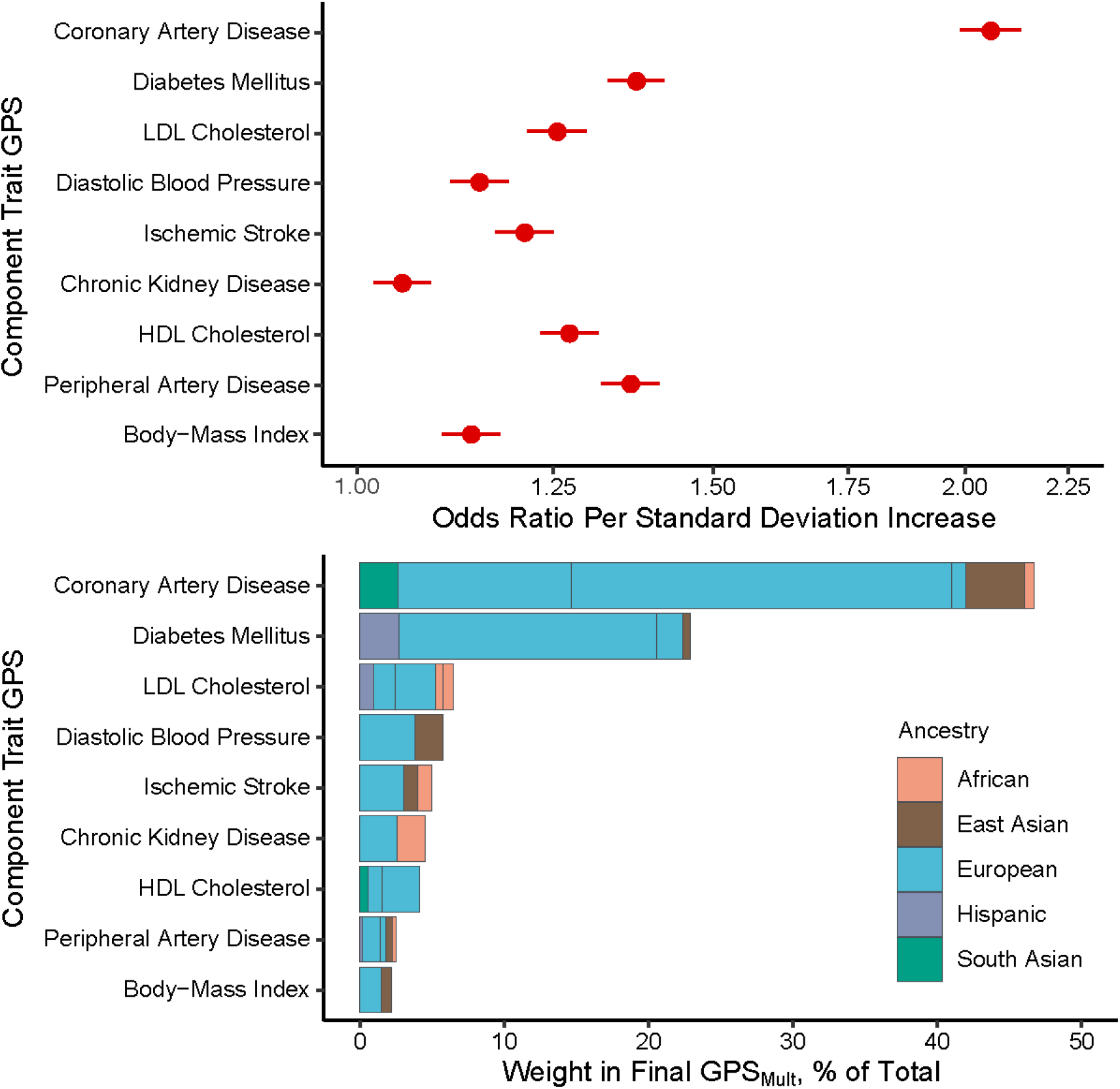
Trait-specific component polygenic score performance and ancestry-specific polygenic score composition of GPS_Mult_ A: The odds ratios for prevalent coronary artery disease (CAD) risk per standard deviation increase of the multi-ancestry, trait-specific layer 1 GPSs were assessed in logistic regression models adjusted for age, sex, genotyping array, and the first ten principal components of ancestry in the same training group of 116,649 UK Biobank European ancestry individuals. B: The contributing weights of each of the ancestry-specific GWAS-based GPS to each of the trait-based layer 1 polygenic scores, color groupings by ancestry of source GWAS, and normalized to 100% to reflect composition in overall GPS_Mult_. Of 58 ancestry- and trait-specific scores that were included in the GPS training analysis, 32 scores significantly contributed to overall prediction in GPS_Mult_ after optimization of score selection with stepAIC and weighting through logistic regression. GPS: genomewide polygenic score; LDL: low-density lipoprotein; HDL: high-density lipoprotein

### Association of GPS_Mult_ with prevalent disease in UK Biobank

The resulting best performing score (GPS_Mult_) demonstrated a strong association with prevalent CAD, with significant improvement from previously published scores. Among 308,264 European ancestry individuals in the hold-out validation dataset, GPS_Mult_ was associated with an odds ratio per standard deviation increase (OR/SD) of 2.14 (95%CI:2.10-2.19) in a model adjusted for age, sex, genotyping array, and the first ten principal components of genetic ancestry, with significant improvement over from prior published scores from the Polygenic Score Catalog without UK Biobank participants in discovery data (Supplementary Table 3).^47^ This corresponded to a Nagelkerke *R*^*2*^ of 0.07 and a logit liability *R*^*2*^ of 0.19 (Supplementary Figure 1). After adjusting for correlated clinical risk factors including systolic and diastolic blood pressure, LDL cholesterol, HDL cholesterol, triglycerides, diabetes, body mass index, and chronic kidney disease, this risk estimate was only modestly attenuated to an OR/SD 2.07 (95% CI 2.02-2.13) (Supplementary Table 4).

The associations between GPS_Mult_ and CAD were largely consistent across studied subgroups, but some evidence of heterogeneity was found when restricting to men (OR/SD 2.20, 95% CI 2.15-2.26, p<0.001) when compared with women (OR/SD 1.94, 95% CI 1.86-2.03, p<0.001), with p-heterogeneity <0.001 (Supplementary Figure 2). Additionally, the association between GPS_Mult_ and CAD was stronger in younger individuals ages 40-54 years (OR/SD 2.17, 95%CI 2.04-2.31, p<0.001) and 55-64 years (OR/SD 2.18, 95%CI 2.11-2.25, p<0.001), when compared with older individuals ages 65-75 years (OR/SD 2.08, 95%CI 2.01-2.15, p<0.001), consistent with recent studies (Supplementary Figure 2).^7,48–50^

GPS_Mult_ showed stronger association with CAD risk when compared with the previously published GPS_2018_^14^ in direct comparison using the same group of individuals for validation. Among individuals with CAD, the median percentile of GPS_Mult_ is significantly higher than that of the GPS_2018_, 75 (IQR 50 - 91) vs 69 (IQR 43 - 88) (Figure 3A). Among individuals of European ancestry, individuals in the bottom and top centile of the polygenic score had a 0.8% and 12.3% prevalence of CAD, respectively, with GPS_2018_, compared with 0.7% and 16.3% prevalence of CAD with and GPS_Mult_ (Figure 3B). Given improved stratification with this newly developed polygenic score, both tails of the score distribution were associated with a greater magnitude of risk when compared with GPS_2018_. With the GPS_2018_, the top 8.3%, 3.1%, and 1.4% of the population had 3-fold, 4-fold, and 5-fold greater odds for CAD relative to the middle quintile of the population, respectively, whereas with the GPS_Mult_, the top 20%, 9.6%, and 4.9% of the population had 3-fold, 4-fold, and 5-fold greater odds for CAD relative to the middle quintile of the population, respectively (Figure 3C, Supplementary Table 5). Conversely, with the GPS_2018_, the bottom 1.7%, 0.5%, and 0.1% of the population had 1/3, 1/4, and 1/5 the odds for CAD relative to the middle quintile of the population, respectively, whereas with the GPS_Mult_, the bottom 13.9%, 1.7%, and 0.2% of the population had 1/3, 1/4, and 1/5 odds for CAD relative to the middle quintile of the population, respectively (Figure 3D).

**Figure 3:**
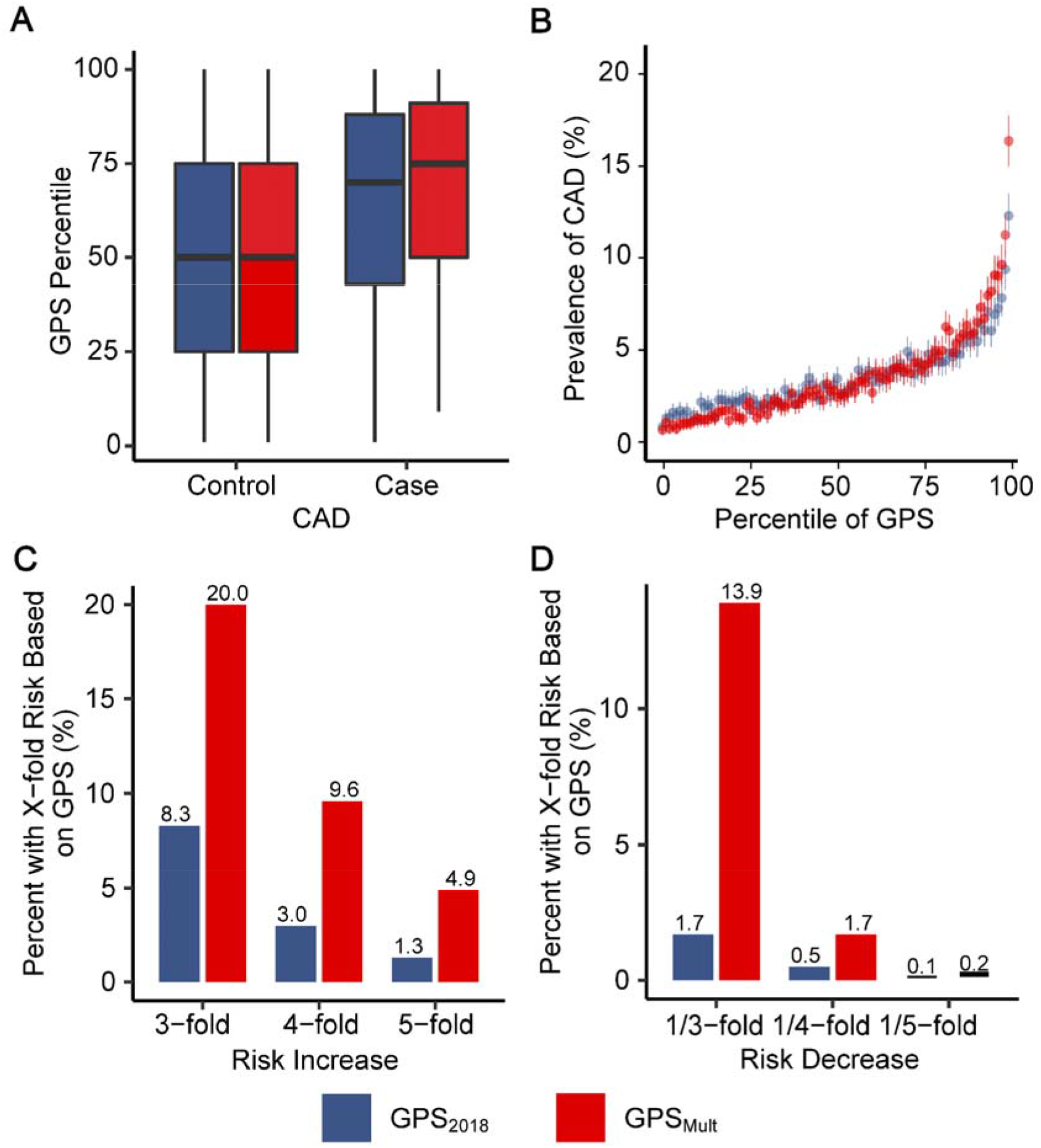
Improvements in polygenic prediction of prevalent coronary artery disease prediction A: Distributions of GPS_2018_ and GPS_Mult_ percentiles across the UK Biobank validation dataset. B: Prevalence of CAD with 95% CI according to 100 groups of the UK Biobank validation dataset binned according to the percentile of the GPS_2018_ and GPS_Mult_. C: Proportion of UK Biobank validation population with 3, 4, and 5-fold increased risk for CAD versus the middle quintile of the population, stratified by GPS. Odds ratio assessed in a logistic regression model adjusted for age, sex, genotyping array, and the first ten principal components of ancestry. D: Proportion of UK Biobank testing population with 1/3, 1/4, and 1/5 risk for CAD versus the middle quintile of the population, stratified by GPS. Odds ratio assessed in a logistic regression model adjusted for age, sex, genotyping array, and the first ten principal components of ancestry. GPS: Genome-wide polygenic score; CAD: coronary artery disease.

### Validation of GPS_Mult_ in external cohorts

GPS_Mult_ was also strongly associated with prevalent CAD in external cohorts, with significant improvement from prior published scores. Published polygenic scores for CAD from the Polygenic Score Catalog and GPS_Mult_ were calculated in an identical group of individuals to facilitate direct comparison within individuals of African and European Ancestry in Million Veteran Program^51^ and South Asian ancestry in Genes & Health^52^ (Figure 4, Supplementary Tables 6-7). For each group, these individuals were not included in published GWAS summary statistics^39,53^ used for GPS_Mult_ derivation. Among 33,096 individuals of African ancestry in the Million Veterans Program, GPS_Mult_ was associated with an OR/SD of 1.24 (95% CI 1.21-1.29, P<0.001) for CAD in a model adjusted for age, sex, genotyping array, and the first ten principal components of genetic ancestry, corresponding in a 73% relative improvement in effect size compared with GPS_2018_ and 39% improvement when compared with the recently published PRS_2022_,^9^ respectively (P=0.008). Similarly, among 124,467 individuals of European ancestry in the Million Veteran Program, GPS_Mult_ was associated with an OR/SD of 1.72 (95% CI 1.69-1.75, P<0.001), corresponding in a 46% and 13.6% relative improvement in effect size compared with GPS_2018_ and PRS_2022_,^9^ respectively (P<0.001). Additionally, among 27,990 individuals of South Asian ancestry in Genes and Health, GPS_Mult_ was associated with an OR/SD of 1.83 (95% CI 1.69-1.99, P<0.02), corresponding to a 113% (P<0.001) and 29% (P=0.02) relative improvement in effect size compared with GPS_2018_ and PRS_2022_, respectively (Figure 4).

**Figure 4:**
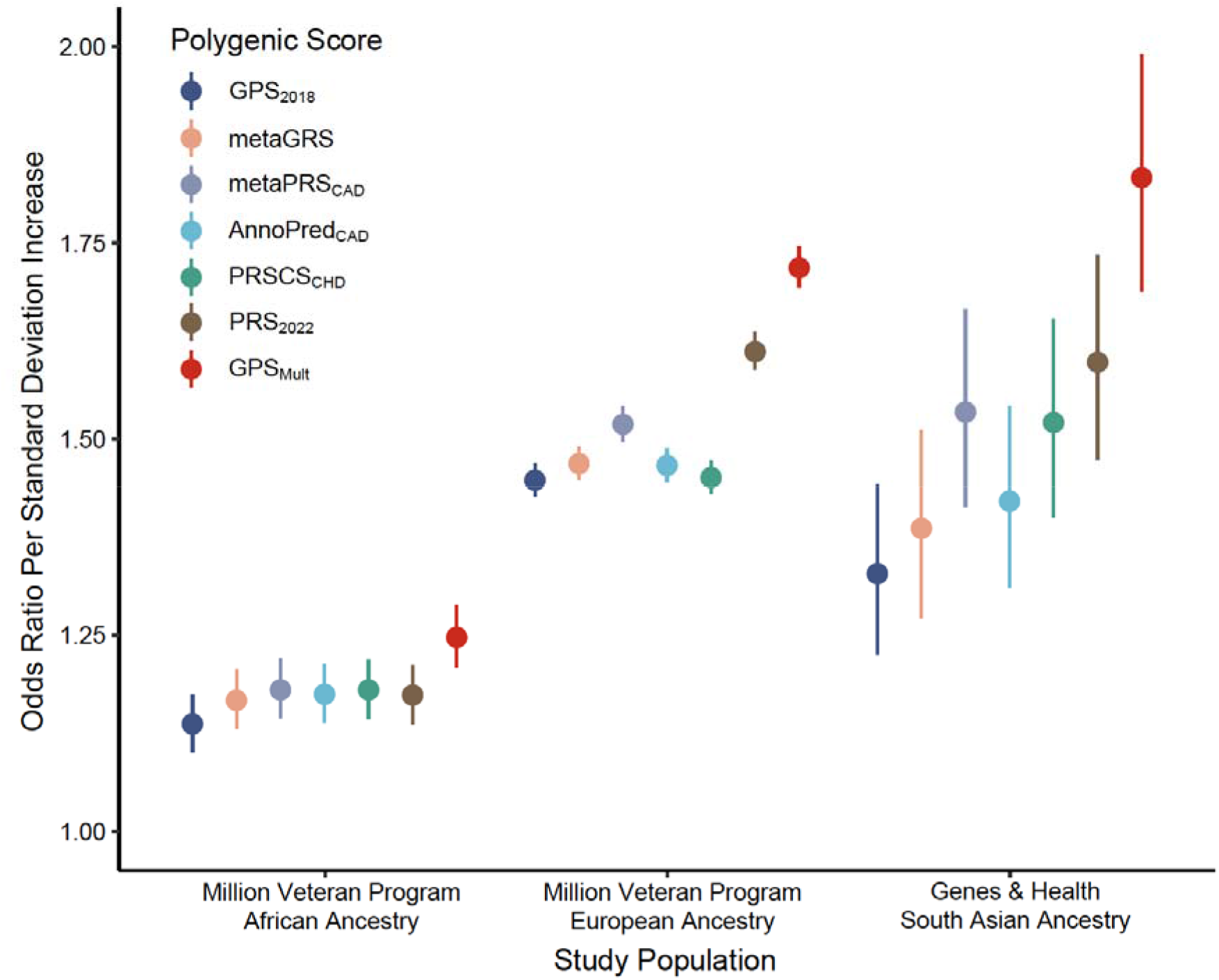
External validation of GPS_Mult_ and benchmarking against published polygenic scores for coronary artery disease across multiple ancestries in Million Veteran Program and Genes & Health studies. The odds ratio for prevalent coronary artery disease (CAD) risk per standard deviation increase of the polygenic score was assessed in a logistic regression model adjusted for age, sex, genotyping array, and the first ten principal components of ancestry in the same group of individuals per cohort: African ancestry individuals in Million Veteran Program; European ancestry individuals in Million Veteran Program; South Asian ancestry individuals in Genes & Health, using high-performing published scores from the Polygenic Score Catalog (GPS_2018_^14^, metaGRS^13^, metaPRS_CAD_^23^, AnnoPred_CAD_^104^, PRSCS_CHD_^75^, and PRS_2022_^9^) and current GPS_Mult_.^47^ Results for these and remaining CAD polygenic scores published in the Polygenic Score Catalog are available in Supplementary Tables 6-7. GPS: Genome-wide polygenic score

### Association of GPS_Mult_ with incident disease in UK Biobank

The GPS_Mult_ was predictive of incident CAD events over median [interquartile range] 12.0 [11.2-12.7] years of follow-up across all four ancestral groups in the UK Biobank. Across the entire UK Biobank study validation population without prior CAD, individuals in the bottom centile of the GPS_Mult_ had a 1.1% incidence of CAD while individuals in the top centile had a 11.7% incidence of CAD. Overall, GPS_Mult_ was associated with a hazard ratio per standard deviation (HR/SD) of 1.73 (95% CI 1.70-1.76, P<0.001), compared with HR 1.49 (95% CI 1.47-1.52, p<0.001) found with GPS_2018_. When stratified by ancestry, risk estimates remained consistent across individuals of East Asian (HR/SD 1.72, 95% CI 1.13-2.60, P=0.011), European (HR/SD 1.75, 95% CI 1.71-1.78, P<0.001), and South Asian (HR/SD 1.62, 95% CI 1.49-1.77, P<0.001) ancestry, but score performance was weakest among individuals of African (HR/SD 1.25, 95% CI 1.07-1.46, p=0.004) ancestry (Figure 5A). Across all individuals in the UK Biobank validation dataset, GPS_Mult_ demonstrated 38% relative improvement in effect size compared with GPS_2018_. Of this, 26% improvement resulted from larger sample size of the primary CARDIOGRAMplusC4D GWAS (excluding UK Biobank participants), 9% improvement from incorporation of multi-ancestry CAD summary statistics, and 3% improvement from leveraging genetic commonalities with CAD risk factors to refine score weighting (Figure 5B). Incorporation of multi-ancestry and multi-trait genetic data resulted in greater relative gains in incident disease prediction for individuals in each ancestry, with improved relative effect sizes of 143%, 71%, 38%, and 23% for individuals of African, East Asian, European, and South Asian ancestry, respectively, compared to GPS_2018_ performance in those groups. These also translated into significant gains in prediction by GPS_Mult_ relative to the initial GPS_2018_ performance in European ancestry, now with improved prediction in African ancestry (relative effect size 0.55 increased from 0.23) (Figure 5B) and performance surpassing the reference score in East Asian ancestry (relative effect size 1.37, increased from 0.80) and South Asian ancestry (relative effect size 1.19, increased from 0.97).

**Figure 5:**
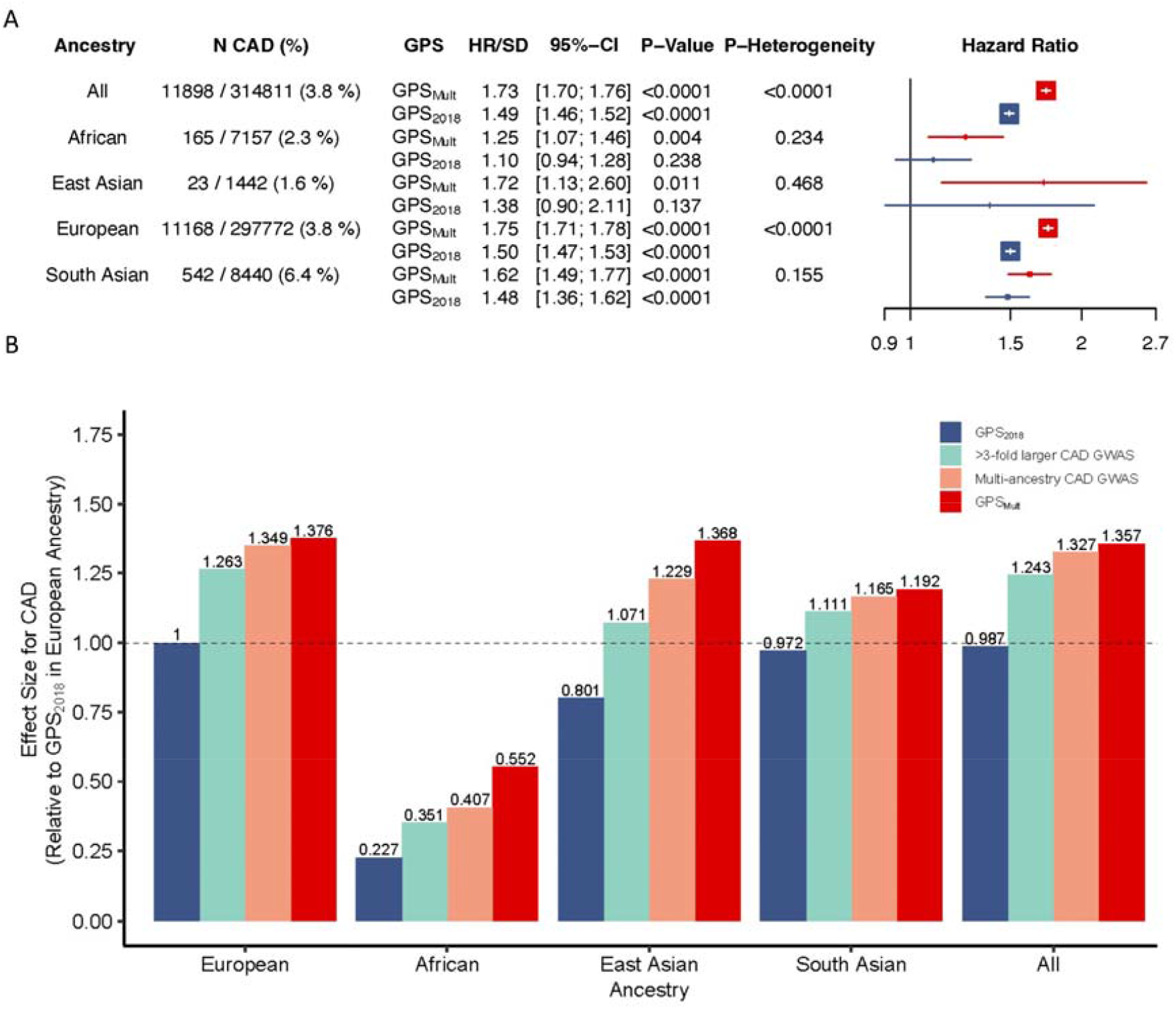
Incident CAD prediction by GPS_Mult_ stratified by ancestry A: Adjusted hazards ratio per standard deviation of the polygenic score with corresponding 95% CIs and P values for coronary artery disease (CAD) by ancestry, stratified by iteration of the version of the polygenic score, calculated from Cox proportional-hazards regression models adjusted for age, sex, genotyping array, and the first ten principal components of ancestry in the UK Biobank validation dataset. GPS_2018_ corresponds to previously published polygenic score for CAD.^14^ B: The score effect sizes relative to the effect size of GPS_2018_ in European ancestry individuals. >3-fold larger CAD GWAS designates metrics for polygenic score generated using summary statistics from the most recent Coronary ARtery DIsease Genomewide Replication and Meta-analysis plus The Coronary Artery Disease Genetics consortium (CARDIOGRAMplusC4D) excluding the UK Biobank, of largely European ancestry. Multi-ancestry CAD GWAS refers to the polygenic score generated by combining ancestry-specific polygenic scores generated using GWAS summary statistics from CARDIOGRAMplusC4D, Genes & Health, Biobank Japan, Million Veteran Program, and FinnGEN biobanks in layer 1. GPS_Mult_ designates polygenic score for CAD designed with summary statistics from multiple ancestries and multiple CAD-related traits in layer 2. *Designates the reference group for calculating relative gain. GPS: Genome-wide polygenic score; CAD: coronary artery disease; GWAS: genome-wide association study.

### Assessment of disease risk in the extremes of the GPS_Mult_ distribution

Additionally, we hypothesized that the GPS_Mult_ could identify individuals in the clinically relevant extreme tails of its distribution. Current cardiovascular disease prevention guidelines recommend statin therapy for individuals with prior coronary artery disease, peripheral artery disease, ischemic stroke, diabetes mellitus, or severe hypercholesterolemia (LDL >=190 mg/dL) to help mitigate their high risk of cardiovascular disease and mortality.^2^ In the high end of GPS_Mult,_ we sought to identify individuals with genetic risk of equivalent magnitude to that of individuals with these clear indications for statin therapy. In prospective analyses of individuals without prior CAD, those within the top 3 percentiles of GPS_Mult_ had equivalent disease risk of incident CAD as the recurrent event risk for an individual who had a CAD event prior to enrollment, adjusting for age and sex (Supplementary Figure 3A).

Furthermore, individuals without peripheral artery disease (PAD) in the top 8% of polygenic score distribution had incident CAD risk equivalent to individuals with prior PAD; individuals without diabetes in the top 21% of polygenic score distribution had incident CAD risk equivalent to individuals with prior diabetes; and individuals without severe hypercholesterolemia (estimated untreated LDL cholesterol ≥ 190 mg/dL) in the top 29% of polygenic score distribution had incident CAD risk equivalent to individuals with prior hypercholesterolemia (Supplementary Figures 3B-D). Conversely, in the low end of the GPS_Mult_ distribution, individuals in the bottom 5 percentiles were associated with a significant reduction in incident CAD risk (HR 0.27, 95%CI 0.21-0.35, P<0.001) when compared with the middle quintile (40-59%). When comparing individuals who smoke and are in the bottom 5 percentiles of GPS_Mult_ with non-smokers in the middle quintile, the associated reduction in the absolute incidence of CAD offsets approximately 60 pack-years of smoking.

Furthermore, individuals in the 5-9th percentiles of GPS_Mult_ had a significant reduction in CAD risk (HR 0.55, 95%CI 0.49-0.62, P<0.001) when compared with the middle quintile. These individuals experienced comparable risk reduction as those individuals carrying variants in *PCSK9* associated lifelong low levels of LDL cholesterol (Supplementary Figure 4).^54,55^

### Modeling of GPS_Mult_ with clinical risk predictors

A risk prediction approach integrating clinical and genetic risk using the American College of Cardiology/American Heart Association Pooled Cohort Equations (PCE),^5^ GPS_Mult_, and their interaction in a single model was used to predict 10-year CAD risk estimates in the UK Biobank validation population. Accounting for the interaction between the polygenic score and clinical risk estimate improves performance beyond the simple addition of the two, with lower GPS_Mult_ weighting with higher PCE estimates (effect size - 0.60, *P*_*interaction*_ *<0*.*001*). This combined model effectively improved risk prediction when compared with PCE alone. When binned by different PCE estimates, this model demonstrated striking stratification of CAD incidence across the GPS_Mult_ distribution, with significant differences observed in ancestry-based subgroups (Figure 6A). The gradient in risk predicted by this model from top to bottom centile was largest in South Asian ancestry individuals with high PCE risk (5.1% to 29.1%), compared with European ancestry individuals (2.6% to 20.6%). When compared with the PCE risk estimate incorporating clinical risk factors alone, integration of the PCE with GPS_Mult_ contributed to significantly higher discrimination and predictive performance across the entire tested population. First, discrimination was assessed in Cox regression models including various covariables using Harrell’s C-Statistic. A gradient in improvement was seen using models using age and sex alone (C-statistic 0.710, 95%CI 0.706 - 0.715), PCE which is inclusive of age and sex (C-statistic 0.739, 95% CI 0.735-0.744), and the model integrating PCE, GPS_Mult_ and their interaction term (C-statistic 0.763, 95%CI 0.759-0.768) (Figure 6B). Similar improvements in C-statistic were observed for models tested in subgroups stratified by ancestry (Supplementary Table 8). Second, categorized net reclassification improvement (NRI) was calculated across the entire study population using a threshold of 7.5% (NRI 0.075) of the predicted 10-year risk of CAD, which is the clinically accepted estimated risk threshold for recommending initiation of statin therapy for prevention of CAD. The risk model combining PCE and GPS_Mult_ resulted in significant improvements in the categorical net reclassification index (NRI = 7.0%, +8.1% for incident cases and -1.1% for non-cases), with GPS_Mult_ resulting in greater up classification of risk largely in individuals who go on to develop disease (Figure 6C). Third, when compared with established risk enhancing factors for CAD risk, categorization within the top 10 percentiles of the GPS_Mult_ distribution corresponded to a significantly higher net reclassification over the use of PCE estimate alone (3.7%) as compared to other risk enhancers like elevated lipoprotein(a) (with NRI 1.3%) (Supplementary Figure 5). Similar results in NRI were observed across other ancestries (Supplementary Table 9). Additionally, similar trends in predictive performance, discrimination, and reclassification were observed with integration of the QRISK score with GPS_Mult_ (Supplementary Tables 8-9).

**Figure 6:**
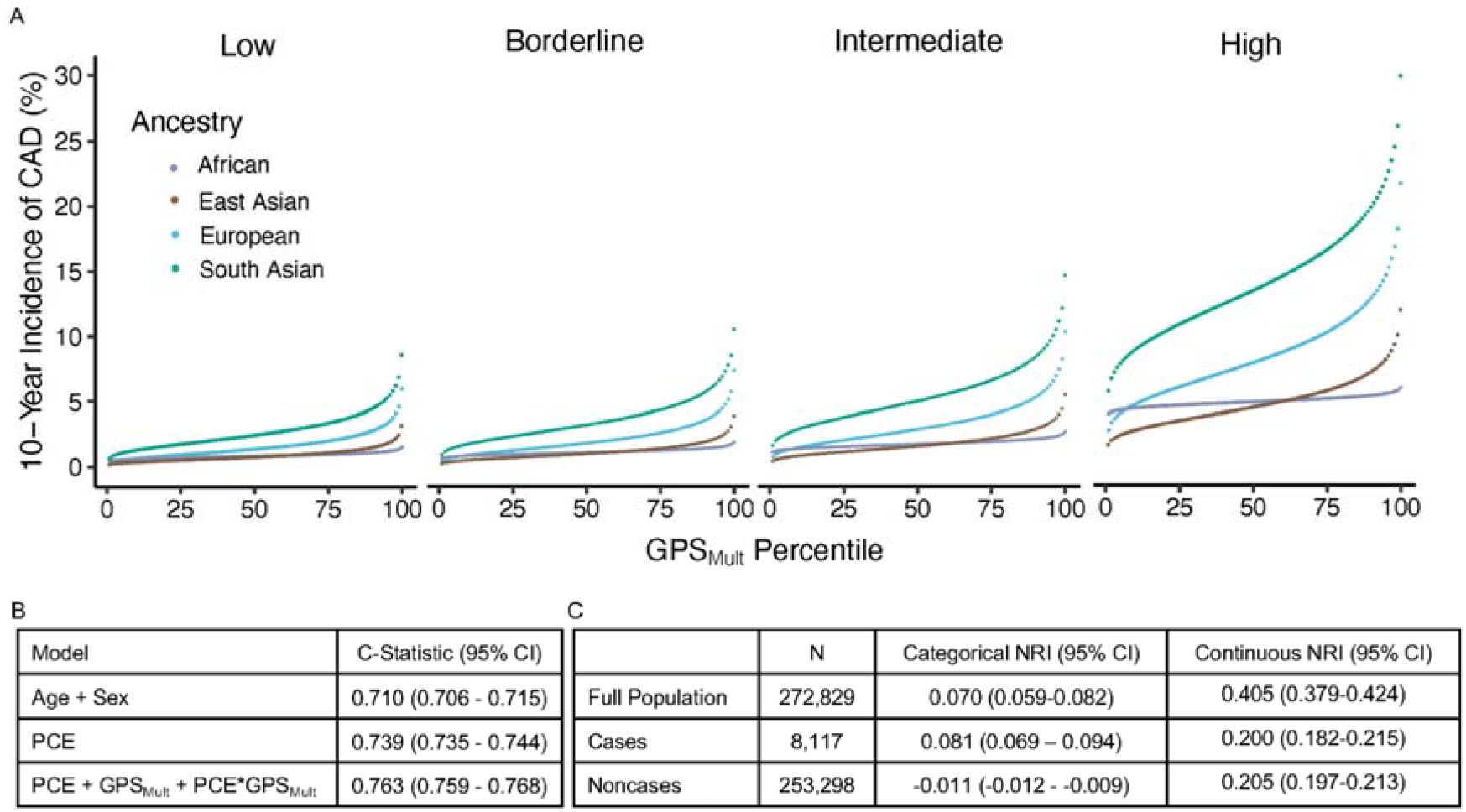
Discrimination and reclassification by a model integrating polygenic and clinical risk for incident CAD A: Cumulative incidence of coronary artery disease (CAD) over 10 years predicted by modeling GPS_Mult_, AHA/ACC Pooled Cohort Equations (PCE) 10-year risk estimate, and their interaction in the UK Biobank validation dataset binned according to the percentile of the GPS_Mult_, grouped by risk categories of the PCE (mean 10-year risk of atherosclerotic cardiovascular disease as low (<5%), borderline (5 to <7.5%), intermediate (≥7.5 to <20%), and high (≥20%)), and stratified by ancestry. B: C-statistics are based on 10-year follow-up events from Cox regression models of listed variables. PCE includes age and sex variables in its risk estimation. C: The improvement in the predictive performance of the addition of the GPS_Mult_ to the PCE was evaluated using continuous and categorised net reclassification improvement (NRI), with a risk probabilities threshold of 7.5% obtained with Kaplan-Meier estimates for a period of 10 years and confidence intervals (95%) obtained from 100-fold bootstrapping. GPS: genome-wide polygenic score.

### Association of GPS_Mult_ with recurrent disease in UK Biobank

In addition to first events, the GPS_Mult_ predicted recurrent CAD events in individuals with prior CAD. GPS_Mult_ was associated with a HR/SD of 1.13 (95% CI 1.08-1.18, P<0.001), comparable to prior studies.^56^ Although a significantly less pronounced effect estimate as compared to prediction of first CAD event, the predictive performance of GPS_Mult_ this context was comparable to that of diastolic blood pressure (HR 1.11, 95%CI 1.06-1.16, P<0.001) and glycated hemoglobin (HR 1.07, 95%CI 1.02-1.12, P<0.001) (Supplementary Figure 6).

## DISCUSSION

A new polygenic score for CAD incorporating multi-ancestry summary statistics from GWAS for CAD and related risk factor traits on a large scale demonstrated significantly improved performance when compared to prior published scores. External validation in fully independent datasets derived from the Million Veterans Program and the Genes & Health studies, confirming enhanced prediction compared to previously published polygenic scores across all studied ancestries. The enhanced predictive capacity of this score was particularly pronounced in the extremes of the score distribution, enabling–in some cases–identification of healthy individuals with risk of CAD equivalent to those with pre-existing disease. When added to risk scores used in current clinical practice, GPS_Mult_ significantly improved discrimination and reclassification relevant to clinically important decision thresholds, such as decision to initiate statin therapy.

This work builds on prior studies in providing a framework for generating the best possible polygenic score for any trait, within the limitations of available GWAS with finite sample sizes and under-representation of diverse populations. The GPS_Mult_ incorporates CAD summary statistics from large non-European ancestry biobanks leading to a total CAD GWAS summary statistics of over 269,000 cases and over 1,178,000 controls, including many-fold larger representation of individuals of non-European ancestries than previously published efforts.^39,52,57–59^ This results in substantial improvements in prediction for individuals of East and South Asian ancestry, reflecting greater representation of summary statistics from Biobank Japan and Genes & Health. However, the majority of improvement in effect size is attributable to use of summary statistics from the largest CAD GWAS to date (CARDIOGRAMplusC4D consortium, excluding UK Biobank participants), particularly in European ancestry individuals.^9^ The modest improvements in prediction observed among individuals of African ancestry are likely due to underrepresentation of this group in GWASs to date.^35^ Due to smaller haplotype blocks observed in individuals of African ancestry, a 4- to 7-fold larger GWAS representation is needed to yield comparable prediction gains.^60^ The additional incorporation of genetic associations with CAD-related risk factors across ancestries into calculating GPS_Mult_ significantly improves prediction beyond using summary statistics from CAD GWAS alone, with impact most notable in individuals of non-European ancestry. This may potentially be due to greater representation of these ancestries in the discovery GWAS for CAD risk factor traits.^61–63^ With these additions, the phenotypic variance explained by GPS_Mult_ for CAD calculated as *R*^*2*^ on the logit-liability scale was 0.19. Although this estimate remains below the estimated SNP heritability for CAD of 0.4 - 0.6, it surpasses the phenotypic variance explained of 0.14 by the largest component GWAS from the CARDIOGRAMplusC4D consortium.^39,64,65^

Improvements in polygenic score performance can help better facilitate clinical decision making. Prospective trials are already underway returning polygenic risk information to patients,^66,67^ and medical societies have begun to provide provisional guidance on their use.^68^ Furthering these goals, GPS_Mult_ is able to better identify individuals at the highest risk for developing incident CAD to potentially guide early preventive interventions.^69,70^ Building on prior work advocating for use of polygenic scores as a risk-enhancing factor to guide decision making regarding statin therapy in individuals at borderline or intermediate CAD risk, the current work more strongly supports use in primary screening across the population to target interventions.^71^ Current cardiovascular prevention guidelines recommend statin initiation for individuals solely based on having any of the following conditions as they portend a high risk of a new atherosclerotic cardiovascular disease event: prior CAD, ischemic stroke, PAD, diabetes, or severe hypercholesterolemia.^2^ This score identified 3% of the population with equivalent risk for a new CAD event as the risk for a recurrent CAD event in individuals who have had prior disease. Similarly, the top 8%, 21%, and 29% of the GPS_Mult_ distribution–despite having no known CAD–had equivalent risk of incident CAD as individuals with prior peripheral artery disease, diabetes mellitus, and severe hypercholesterolemia, respectively. Because all three of these designations are currently clinical indications for statin therapy, a high GPS_Mult_ could be employed to identify additional individuals for cholesterol-lowering therapies as an adjunct to current guidelines.

Furthermore, given the GPS_Mult_’s ability to identify these individuals with the highest propensity for developing CAD, these scores could be employed to enrich for high genetic risk individuals in CAD prevention trials to maximize event rates and minimize drug trial costs.^72^ The GPS_Mult_ could also be employed to identify the individuals with the highest risk of recurrent events for targeted, otherwise costly therapies which have been shown to be beneficial in this population.^73,74^ Additionally, GPS_Mult_ also identifies individuals in the lower end of genetic risk who are seemingly protected from CAD with similar risk reduction as that of carriers of variants in the *PCSK9* gene leading to lifelong reductions on low-density lipoprotein cholesterol.^54,55^

Furthermore, a risk model incorporating polygenic risk with the PCE estimated risk is applied to individuals across different ancestries to demonstrate improved predictive performance.^43,44^ This improved performance illustrates the potential for an integrated absolute risk prediction model.^43–45,75^ For example, this model is particularly useful in differentiating risk in the high-risk South Asian ancestry population, where traditional clinical risk estimators often fail to capture the increased risk associated with this ancestry.^4^ The integration of the GPS_Mult_ with PCE builds on prior efforts which demonstrated improvement in model discrimination by now showing nearly identical improvement in C-statistic (0.03) in between models incorporating i) age and sex, ii) PCE alone, and iii) combined genetic and clinical risk across the population.^7,76,77^ However measures of C-statistic alone are not optimal or fully comprehensive in evaluating models that predict future risk.^78^ GPS_Mult_ demonstrates nearly three-fold greater net reclassification of CAD cases/noncases when added to the PCE 10-year risk assessment to guide statin initiation as compared with established ‘risk enhancing factors.’^79^ Further work is needed to incorporate additional risk factors. To aid in future model calibration efforts, there is a need for population-level disease incidence and mortality data disaggregated by ancestral sub-groups.^67^

These results should be interpreted within the context of limitations. Polygenic scores were developed and validated in individuals of European ancestry and then externally validated in non-European ancestry populations, and this may be partially contributing to poorer predictive performance in these groups. These results underscore the need for larger and more representative GWAS studies. UK Biobank participants were recruited at age 40-69 years, raising the possibility of survivorship or selection bias that limits generalizability to younger patients, however recent studies have demonstrated reliable performance of GPS in younger age groups.^8^ All UK Biobank disease endpoints were similarly ascertained through participant self-report, diagnosis codes from inpatient admissions, national procedure, and death registries. Participants in research studies tend to be healthier than the general population^80^ — recalibration of disease risk models for a given target population may be needed prior to clinical deployment.

In conclusion, incorporating GWAS data for CAD and related traits from multiple ancestries on a large-scale leads to significantly improved performance of GPS_Mult_ in external validation among diverse ancestry populations when compared with previously published scores. This approach is generalizable to all traits and results in a polygenic score that is able to better identify individuals at the highest and lowest ends of risk, significantly reclassifies risk beyond clinical risk estimators, and has the potential to advance clinical decision making.

## METHODS

### Data availability

All data are made available from the UK Biobank to researchers from universities and other institutions with genuine research inquiries following institutional review board and UK Biobank approval. This research was conducted using the UK Biobank resource under Application Number 7089 and secondary data use was approved by the Mass General Brigham institutional review board. Summary statistics from Biobank Japan are available at http://jenger.riken.jp/en/result. Summary statistics for the Coronary ARtery DIsease Genomewide Replication and Meta-analysis plus The Coronary Artery Disease Genetics consortium (CARDIoGRAMplusC4D) study are available at http://www.cardiogramplusc4d.org. Summary statistics from FinnGen are available at https://www.finngen.fi/en/access_results. Summary statistics from Genes & Health are available at https://www.genesandhealth.org/research/scientific-data-downloads. Summary statistics from the Million Veteran Program are available in dbGaP (accession number phs001672). The full GPS_Mult_ weights will be made available in the Polygenic Score Catalog.

### Study populations

The UK Biobank is a prospective cohort study that enrolled over 500,000 individuals between the ages of 40 and 69 years between 2006 and 2010.^46,81^ A detailed questionnaire completed by UK Biobank participants at enrolment assessed self-report of ancestry, lifestyle factors, including smoking. Anthropometric measurements including body-mass index were measured at the initial enrollment visit. Biomarkers including serum lipid concentrations and renal function markers were assessed at time of enrolment as part of the study protocol. Diagnoses of peripheral artery disease (PAD), diabetes, and hypertension were determined based on self-report or hospitalization records confirming a clinical diagnosis, as previously described.^4,82^

Participants within the Million Veteran Program were recruited from more than 75 Veteran Affairs Medical Centers nationwide since 2011, with >885,000 individuals currently enrolled.^51^ Each participant has consented to linkage to their electronic medical record, wherein ICD9/10 diagnosis codes, Current Procedural Terminology (CPT) codes, clinical laboratory measurements, and reports of diagnostic imaging modalities are available. Participants were also asked to complete baseline and lifestyle questionnaires to further augment data contained in the electronic health record.

Genes & Health is a UK-based cohort of over 48,000 British Pakistani and Bangladeshi individuals recruited and consented for lifelong electronic health record access and genetic analysis.^52^ Medical records are linked to ICD10, OPCS, and SNOMED diagnosis and procedural codes across inpatient and hospital settings as well as clinical laboratory measurements, and a baseline questionnaire.

### Clinical endpoints

Ascertainment of CAD at enrollment in the UK Biobank was based on self-report, hospitalization records or death registry confirming diagnosis of myocardial infarction or its acute complications, or a coronary revascularization procedure (coronary artery bypass graft surgery or percutaneous angioplasty/stent placement), as previously described.^82,83^ The earliest date at which the diagnosis was ascertained was considered as the diagnosis date. For individuals with CAD prior to enrollment, recurrence of CAD was determined based on diagnosis of a myocardial infarction or revascularization in the follow-up period, as previously described.^84^

Within the Million Veteran Program, ICD9, ICD10, and CPT codes from both inpatient and outpatient encounters were used to curate and classify CAD cases based on having a myocardial infarction or undergoing revascularization, identified as subjects with at least two codes (of any category) that occurred on distinct dates within a 12 month window, as previously described.^39^ Incident cases were identified as those with the first of the two qualifying codes occurring after enrollment. The remaining CAD cases, including through self-report, were considered prevalent.

In the Genes & Health study, ICD10 and SNOMED codes from the linked electronic health record were used to classify CAD cases defined as myocardial infarction or revascularization based on first diagnosis date, as described elsewhere.^53^ Prevalent cases were defined as events prior to enrollment while events occurring after enrollment were designated as incident disease.

### GPS construction

Summary statistics from recent CAD GWAS studies (Genes & Health, FinnGen, Million Veterans Program, Biobank Japan, and CARDIOGRAMplusC4D excluding UK Biobank samples) conducted in individuals of diverse ancestries were used to determine primary CAD score weights (Supplementary Table 1).^9,39,52,57,58^ UK Biobank participants were not included among these discovery cohorts to preserve them as an independent hold-out dataset for training and validation of the GPS_Mult_ (Supplementary Table 2). Ancestry-specific linkage disequilibrium reference panels were extracted from the 1000 Genomes Project phase 3 data to match with the ancestry for the discovery GWAS, and only unrelated samples were used.^85^ GPS_Mult_ construction was comprised in a two layer process, with layer 1 consisting of combining multiple polygenic scores derived from different ancestry-specific GWAS data for each trait, and layer 2 consisting of combining the multi-ancestry CAD polygenic score with similarly constructed multi-ancestry CAD-related trait scores predicting CAD (Figure 1) to generate GPS_Mult_.

Separate GPS were constructed for each ancestry-stratified CAD GWAS using the LDPred2 method, which is a Bayesian approach to calculate a posterior mean effect for all variants based on an effect size in the prior GWAS and subsequent shrinkage based on linkage disequilibrium.^86^ Only HapMap3 variants – a set of >1.4 million variants compiled by the International HapMap Project which capture common patterns of variation in a variety of human populations – were included for score calculation.^87^ The default parameters used in the LDPred2 method included the proportion of variants to be causal (cut-offs of *p*=1.0×10^−04^, 1.8×10^−04^, 3.2×10^−04^, 5.6×10^−04^, 1.0×10^−03^, 1.8×10^−03^, 3.2×10^−03^, 5.6×10^−03^, 1.0×10^−02^, 1.8×10^−02^, 3.2×10^−02^, 5.6×10^−02^, 1.0×10^−01^, 1.8×10^−01^, 3.2×10^−01^, 5.6×10^−01^ and 1), the scale of heritability (s=0.7,1 and 1.4), and whether or not a sparse LD matrix was applied.^14,86,88^ Combinations of these parameters resulted in 102 candidate GPSs for each set of ancestry-stratified GWAS summary statistics. The best GPS was selected among these candidates by assessing their performance in predicting prevalent CAD in an independent 116,649 individuals of White British ancestry from UK Biobank (this data set was used in all the score selection procedures thereafter and same group of individuals used to train previously published score GPS_2018_).^14^ For selecting the best combination of CAD GPS scores from each ancestry-specific CAD GWAS for mixing, the discriminative capacities (Akaike information criterion,

AIC) of these GPS combinations for predicting CAD were assessed using the stepAIC function from R MASS package.^89^ A logistic regression model was used to estimate the mixing weights for each individual ancestry-specific GPS. These GPSs were then linearly combined together into a single CAD_GPS_ score (layer 1, Figure 1). Similar procedures were followed for other atherosclerotic diseases (ischemic stroke, PAD)^61,90^ and risk factor traits – LDL cholesterol, HDL cholesterol, triglycerides^62,79^, diabetes^91^, systolic blood pressure^92^, diastolic blood pressure, chronic kidney disease^63^, body-mass index^93^ (Supplementary Table 1, Figure 1).

Then, these multi-ancestry, trait-specific GPSs were linearly combined with the multi-ancestry CAD GPS (from layer 1) to generate the final GPS_Mult_ (layer 2). Just as for layer 1, the discriminative capacities (AIC) of these GPS combinations for predicting CAD were assessed to identify the best combination of trait-level scores for mixing.^89^ A logistic regression model was used to estimate the mixing weights for each individual ancestry-specific GPS using the stepAIC function as described above. These GPSs were then linearly combined together into a single GPS_Mult_ score (layer 2, Figure 1). Of 58 GWAS- and ancestry-specific GPS that went through layers 1 and 2 of selection and mixing, 32 contributed to the final GPS_Mult_, incorporating GWAS summary statistics from multiple ancestries and multiple CAD-related traits (Figure 2). LDPred2 parameters selected for each score, whether the score survived after feature selection, and mixing weights from layers 1 and 2 are listed in Supplementary Table 1.

### GPS validation

The GPS_Mult_ was compared to previously published polygenic scores for CAD. The variant effect sizes were downloaded from PGS Catalog and calculated in the same UK Biobank validation dataset of 308,264 European ancestry individuals for direct comparison.^13–15,47,59,70,75,76,94–105^ See Supplementary Table 3-5 for score accession numbers and performance metrics. The validation datasets were composed of UK Biobank participants separate from those used to train the GPS_Mult_. These individuals underwent genotyping using the UK BiLEVE Axiom Array or UK Biobank Axiom Array, containing over 800,000 variants spanning the genome.^46^ Imputation was performed using the Haplotype Reference Consortium resource, the UK10K panel, and the 1000 Genomes panel.^85,106,107^ We identified a subset of 488,243 participants with genotyping array data. After additional exclusion of 45,602 individuals for high heterozygosity or genotype missing rates, discordant reported versus genotypic sex, putative sex chromosome aneuploidy, excess relatedness (third-degree relative or closer), withdrawal of informed consent derived centrally, or unreported ancestry and 116,650 individuals used for score training, 325,991 individuals (54.3% female, 2.2% African, 0.4% East Asian, 92.0% European, and 2.7% South Asian) were included in the validation cohort for subsequent analyses.

Among Million Veteran Program participants, 157,563 individuals not included in the previously published CAD GWAS^39^ were included and comprised of 33,096 (21%) individuals of African ancestry and 124,467 (79%) individuals of European ancestry (Supplementary Table 2). Individuals were genotyped using the Affymetrix Axiom array and imputed to the TOPMed reference panel. Variants and sample quality control were previously described.^108^

Within the Genes & Health study, individuals not included in the previously published CAD GWAS^53^ were included and comprised 16,874 participants of South Asian ancestry (Supplementary Table 2). These individuals underwent genotyping using the Illumina Infinium Global Screening Array v3 and imputed using the GenomeAsia pilot reference panel. Variants with low call rate (<0.99), rare variants with minor allele frequency (MAF)□<□1%, and variants that failed the Hardy–Weinberg test (*p*□<□1□×□10^−6^) in a subset of samples with low level of autozygosity were removed.

Across all cohorts, individuals were analyzed in distinct self-identified groups of African, East Asian, European, and South Asian ancestries. The generated polygenic scores were residualized for the first ten principal components of genetic ancestry and then scaled to a mean of 0 and standard deviation of 1 for each ancestral group.

### Statistical analysis

Comparison of baseline characteristics between individuals with high or average genetic risk based on polygenic score was performed with the Chi-squared test for categorical variables, analysis of variance (ANOVA) for a subset of continuous variables with normal distributions, and Mann-Whitney U test for continuous variables with nonparametric distributions. Individuals with a given magnitude of increased risk were identified by comparing progressively higher percentile cut-offs to the middle quintile population in a logistic regression model predicting disease status and adjusted for baseline model covariates. Individuals were next binned into 100 groupings according to percentile of the GPS_Mult_ and the unadjusted prevalence of CAD within each bin was determined.

Risk for prevalent disease was calculated using logistic regression models, including baseline model covariates defined as enrollment age, sex, genotyping array, and the first 10 principal components of genetic ancestry. Risk for incident CAD was calculated using Cox proportional-hazards regression models, including baseline model covariates. The proportion of phenotypic variance explained by the polygenic score or risk factor of interest on the observed scale was calculated using the Nagelkerke’s pseudo-*R*^2^ metric– where *R*^2^ was calculated for the full model inclusive of the variable of interest plus the baseline model covariates minus *R*^2^ for the baseline model covariates alone. The proportion of phenotypic variance explained on the liability scale was similarly calculated using the logit liability *R*^2^ metric, as described elsewhere.^109^

To determine the polygenic risk equivalent of a CAD event comparable to risk experienced by those with prior CAD, a model was constructed comparing three groups and monitored for a CAD event in the follow-up period: individuals with prior CAD, individuals without prior CAD in different groupings of the top distribution of GPS_Mult_ (high GPS_Mult_) and the remaining individuals without prior CAD. Sequentially lower percentile cut-offs for this high GPS_Mult_ group were tested to find the grouping with equivalent risk increase for CAD as those with prior CAD. This analysis was repeated for diabetes mellitus, PAD, and severe hypercholesterolemia (LDL cholesterol ≥190 mg/dL). In the lower tail of GPS_Mult_, the risk for incident CAD was calculated in individuals in the bottom 5 percentiles or 5-9th percentiles of GPS_Mult_ relative to those in the middle quintile, using Cox proportional hazards regression models including baseline model covariates. The prevalence of CAD among individuals in the bottom 5 percentiles of GPS_Mult_ was calculated, stratified by 20 pack-years smoking increments and compared with the prevalence of CAD in non-smokers in the middle 40-59 percentiles to estimate equivalent offset risk.

Cox proportional hazards models were used to estimate hazard ratios for incident CAD in the UK Biobank, with covariates of the first 10 principal components. In model 1, only age and sex were modeled with the covariates. In model 2, only the clinical risk estimator – ACC/AHA Pooled Cohort Equations (PCE)^5^ or QRISK3^6^ – was modeled with the covariates. In model 3, GPS_Mult_, clinical risk estimator, and the interaction term of GPS_Mult_ with the clinical risk estimator, and the first 10 principal components of genetic ancestry are modeled.

The 10-year incidence of CAD for individuals grouped by GPS_Mult_ percentile and stratified by ancestry group was quantified using model 3 standardized to four PCE risk levels (mean 10-year risk of atherosclerotic cardiovascular disease as low (<5%), borderline (5 to <7.5%), intermediate (≥7.5 to <20%), and high (≥20%)) and the means of each of the covariates. The discrimination of each of these predictive models was assessed using Harrell’s C-statistic. The improvement in predictive performance of the addition of the GPS_Mult_ to the PCE or QRISK3 was evaluated using continuous and categorized net reclassification improvement (NRI), with a risk probability threshold of 7.5% obtained with Kaplan-Meier estimates for a period of 10 years and 95% confidence intervals obtained from 100-fold bootstrapping. All statistical analyses were performed with the use of R software, versions 3.5 and 3.6 (R Project for Statistical Computing).

## Data Availability

All data are made available from the UK Biobank to researchers from universities and other institutions with genuine research inquiries following institutional review board and UK Biobank approval. This research was conducted using the UK Biobank resource under Application Number 7089 and secondary data use was approved by the Mass General Brigham institutional review board. Summary statistics from Biobank Japan are available at http://jenger.riken.jp/en/result. Summary statistics for the Coronary ARtery DIsease Genome wide Replication and Meta-analysis plus The Coronary Artery Disease Genetics consortium (CARDIoGRAMplusC4D) study are available at http://www.cardiogramplusc4d.org. Summary statistics from FinnGen are available at https://www.finngen.fi/en/access_results. Summary statistics from Genes & Health are available at https://www.genesandhealth.org/research/scientific-data-downloads. Summary statistics from the Million Veteran Program are available in dbGaP (accession number phs001672). The full GPSMult weights will be available in the Polygenic Score Catalog.

## ACKNOWLEDGEMENTS

This work was supported by the KL2/Catalyst Medical Research Investigator Training award from Harvard Catalyst (to A.P.P. and K.G.A.); the Sarnoff Cardiovascular Research Foundation Fellowship (to S.A.); grants 1K08HL153937 (to K.G.A.),1K08HL161448 (to A.C.F.), R01HL1427 (to P.N.), R01HL148565 (to P.N.), R01HL148050 (to P.N.), 1RO1HL092577 (to P.T.E.), 1R01HL157635 (to P.T.E.), and 1R01HL157635 (to P.T.E.) from the National Heart, Lung, and Blood Institute; grant 862032 (to K.A.) and grants 18SFRN34110082 (to P.T.E.), 17IFUNP3384001 (to .G.A.) from the American Heart Association; grant MAESTRIA 965286 from the European Union (to P.T.E.); grants 1K08HG010155 and 1U01HG011719 from the National Human Genome Research Institute (to A.P.P., P.N., and A.V.K.); a Hassenfeld Scholar Award from Massachusetts General Hospital (to P.N. and A.V.K.); a Merkin Institute Fellowship from the Broad Institute of MIT and Harvard (to A.V.K.). This research has been conducted using the UK Biobank Resource, and we thank the volunteers participating. This research is based on data from the Million Veteran Program, Office of Research and Development, Veterans Health Administration, and was supported by Veterans Administration awards I01–01BX003362, I01-BX004821 (P.S.T), I01-BX003340 (P.W.F.W) and VA HSR RES 13–457 (VA Informatics and Computing Infrastructure). The content of this manuscript does not represent the views of the Department of Veterans Affairs or the United States Government. Genes & Health is/has recently been core-funded by Wellcome (WT102627, WT210561), the Medical Research Council (UK) (M009017, MR/X009777/1, MR/X009920/1), Higher Education Funding Council for England Catalyst, Barts Charity (845/1796), Health Data Research UK (for London substantive site), and research delivery support from the NHS National Institute for Health Research Clinical Research Network (North Thames). Genes & Health is/has recently been funded by Alnylam Pharmaceuticals, Genomics PLC; and a Life Sciences Industry Consortium of Astra Zeneca PLC, Bristol-Myers Squibb Company, GlaxoSmithKline Research and Development Limited, Maze Therapeutics Inc, Merck Sharp & Dohme LLC, Novo Nordisk A/S, Pfizer Inc, Takeda Development Centre Americas Inc. We thank Social Action for Health, Centre of The Cell, members of our Community Advisory Group, and staff who have recruited and collected data from volunteers. We thank the NIHR National Biosample Centre (UK Biocentre), the Social Genetic & Developmental Psychiatry Centre (King’s College London), Wellcome Sanger Institute, and Broad Institute for sample processing, genotyping, sequencing and variant annotation. We thank: Barts Health NHS Trust, NHS Clinical Commissioning Groups (City and Hackney, Waltham Forest, Tower Hamlets, Newham, Redbridge, Havering, Barking and Dagenham), East London NHS Foundation Trust, Bradford Teaching Hospitals NHS Foundation Trust, Public Health England (especially David Wyllie), Discovery Data Service/Endeavour Health Charitable Trust (especially David Stables), NHS Digital - for GDPR-compliant data sharing backed by individual written informed consent. Most of all we thank all of the volunteers participating in Genes & Health.

