## Supplementary Appendix for "Polygenic score informed by genome-wide association studies of multiple ancestries and related traits improves risk prediction for coronary artery disease"

### Table of Contents

***Supplementary Tables:***

Table 1: $GPS_{Mult}$ inputs and parameters

Table 2: Summary of $GPS_{Mult}$ training and validation datasets

Table 3: Performance of polygenic scorers for CAD from Polygenic Score Catalog in the UK Biobank Study

Table 4: Baseline risk factor distributions and correlations with $GPS_{Mult}$ in the UK Biobank Study

Table 5: Baseline characteristics according to high $GPS_{Mult}$ in the UK Biobank Study

Table 6: Performance of polygenic scorers for CAD from Polygenic Score Catalog in the Million Veteran Program Study

Table 7: Performance of polygenic scorers for CAD from Polygenic Score Catalog in the Genes & Health Study

Table 8: Model C-statistics by ancestry in the UK Biobank Study

Table 9: Model net reclassification by ancestry in the UK Biobank Study

***Supplementary Figures:***

Figure 1: Sequential improvements in $R^2$ with $GPS_{Mult}$

Figure 2: $GPS_{Mult}$ performance by sex and age subgroups

Figure 3: Equivalents of increased risk for incident coronary artery disease event identified by high $GPS_{Mult}$

Figure 4: Decrease in risk for incident coronary artery disease identified by low $GPS_{Mult}$

Figure 5: NRI for $GPS_{Mult}$ and CAD risk enhancers over PCE 10-year risk estimates

Figure 6: Association of $GPS_{Mult}$ and risk factors with incident and recurrent coronary artery disease events

***References***

**Table 1:** $GPS_{Mult}$ inputs and parameters

| Trait | Name | Dominant Ancestry | N Cases | N Controls | ρ | $h^2$ scale | Is sparse LD | Train OR/SD | Layer 1 Mixing weight | Layer 2 Mixing weight | Final Weight | Reference |
|---|---|---|---|---|---|---|---|---|---|---|---|---|
| | CARDIO-GRAM plusC4D no UKBB | EUR | 86847 | 417789 | 0.018 | 1 | F | 1.92 | 0.51 | | 0.263 | 1 |
| | BBJ | EAS | 29319 | 183134 | 0.01 | 0.7 | F | 1.43 | 0.08 | | 0.041 | 2 |
| CAD | Genes & Health | SAS | 1110 | 20898 | 0.0056 | 1.4 | F | 1.11 | 0.05 | 0.67 | 0.026 | 3 |
| | FinnGen | FIN | 33628 | 275526 | 0.0032 | 1.4 | F | 1.46 | 0.02 | | 0.010 | 4 |
| | MVP | EUR | 95151 | 197287 | 0.018 | 0.7 | F | 1.72 | 0.23 | | 0.120 | 5 |
| | MVP | AFR | 17202 | 59507 | 0.018 | 1.4 | F | 1.10 | 0.01 | | 0.006 | 5 |
| | MVP | HISP | 6378 | 24270 | 0.0018 | 0.7 | F | 1.16 | 0.00 | | 0.000 | 5 |
| | GIANT | EUR | 339224 | | 1 | 1 | T | 1.12 | 0.11 | | 0.014 | 6 |
| BMI | BBJ | EAS | 163835 | | 0.32 | 0.7 | F | 1.06 | 0.05 | 0.03 | 0.007 | 2 |
| | MVP | EUR | 249262 | | 0.032 | 1.4 | F | 1.11 | 0.12 | | 0.038 | 7 |
| DBP | BBJ | EAS | 145515 | | 0.01 | 0.7 | F | 1.07 | 0.06 | 0.08 | 0.019 | 2 |
| | MVP | EUR | 249262 | | 0.032 | 0.7 | F | 1.19 | 0.19 | | 0.000 | 7 |
| SBP | BBJ | EAS | 145505 | | 0.018 | 1.4 | F | 1.09 | 0.05 | 0.00 | 0.000 | 2 |
| | MVP | EUR | 148726 | 965732 | 1 | 1.4 | T | 1.31 | 0.29 | | 0.179 | 8 |
| | MVP | AFR | 24646 | 31446 | 0.018 | 0.7 | F | 1.06 | 0.00 | | 0.000 | 8 |
| DM | MVP | HISP | 8,616 | 11,829 | 0.032 | 1.4 | T | 1.09 | 0.04 | 0.33 | 0.027 | 8 |

| | | | | | | | | | | | | |
|---|---|---|---|---|---|---|---|---|---|---|---|---|
| | AGEN T2D | EAS | 77418 | 356122 | 0.56 | 1 | T | 1.14 | 0.01 | | 0.005 | [9] |
| | FinnGen | FIN | 49303 | 255466 | 0.032 | 1.4 | F | 1.16 | 0.03 | | 0.018 | [4] |
| | Genes & Health | SAS | 9044 | 12066 | 0.0056 | 1.4 | F | 1.05 | 0.00 | | 0.000 | [10] |
| | GLGC | EUR | 1320016 | | 0.1 | 0.7 | T | 1.22 | 0.26 | | 0.028 | [11] |
| | GLGC | AFR | 99432 | | 0.01 | 0.7 | T | 1.15 | 0.07 | | 0.007 | [11] |
| | GLGC | SAS | 40963 | | 0.00032 | 1.4 | T | 1.15 | 0.00 | | 0.000 | [11] |
| | MVP | EUR | 215551 | | 0.01 | 0.7 | T | 1.17 | 0.14 | | 0.015 | [12] |
| | MVP | AFR | 57332 | | 0.0056 | 0.7 | T | 1.13 | 0.05 | | 0.005 | [12] |
| | MVP | HISP | 24742 | | 0.0018 | 0.7 | T | 1.18 | 0.09 | | 0.009 | [12] |
| LDL-C | BBJ | EAS | 72866 | | 0.0032 | 0.7 | T | 1.13 | 0.00 | 0.09 | 0.000 | [2] |
| | GLGC | EUR | 1320016 | | 1 | 0.7 | T | 1.24 | 0.07 | | 0.009 | [11] |
| | GLGC | AFR | 99432 | | 0.32 | 1.4 | F | 1.08 | 0.00 | | 0.000 | [11] |
| | GLGC | SAS | 40963 | | 0.001 | 1 | T | 1.05 | 0.04 | | 0.006 | [11] |
| | MVP | EUR | 215551 | | 0.32 | 1 | F | 1.20 | 0.20 | | 0.026 | [12] |
| | MVP | AFR | 57332 | | 0.0056 | 1 | F | 1.07 | 0.00 | | 0.000 | [12] |
| | MVP | HISP | 23946 | | 0.056 | 1.4 | F | 1.08 | 0.00 | | 0.000 | [12] |
| HDL-C | BBJ | EAS | 74970 | | 0.32 | 0.7 | F | 1.06 | 0.00 | 0.06 | 0.000 | [2] |
| | GLGC | EUR | 1320016 | | 0.18 | 0.7 | T | 1.20 | 0.13 | | 0.000 | [11] |
| TC | GLGC | AFR | 99432 | | 0.01 | 0.7 | T | 1.13 | 0.00 | 0.00 | 0.000 | [11] |

| | | | | | | | | | | | | |
|---|---|---|---|---|---|---|---|---|---|---|---|---|
| | GLGC | SAS | 40963 | | 0.00018 | 1.4 | F | 1.14 | 0.00 | | 0.000 | 11 |
| | MVP | EUR | 215551 | | 0.01 | 0.7 | T | 1.16 | 0.00 | | 0.000 | 12 |
| | MVP | AFR | 57332 | | 0.0056 | 0.7 | F | 1.11 | 0.00 | | 0.000 | 12 |
| | MVP | HISP | 24743 | | 0.001 | 0.7 | F | 1.17 | 0.07 | | 0.000 | 12 |
| | BBJ | EAS | 135808 | | 0.001 | 0.7 | F | 1.12 | 0.04 | | 0.000 | 2 |
| | GLGC | EUR | 1320016 | | 1 | 0.7 | T | 1.24 | 0.24 | | 0.000 | 11 |
| | GLGC | AFR | 99432 | | 0.018 | 1.4 | F | 1.09 | 0.00 | | 0.000 | 11 |
| | GLGC | SAS | 40963 | | 0.01 | 0.7 | F | 1.09 | 0.00 | | 0.000 | 11 |
| | MVP | EUR | 215551 | | 0.18 | 1 | F | 1.19 | 0.00 | | 0.000 | 12 |
| | MVP | AFR | 57332 | | 0.032 | 0.7 | F | 1.07 | 0.00 | | 0.000 | 12 |
| | MVP | HISP | 24063 | | 0.018 | 1.4 | F | 1.08 | 0.00 | | 0.000 | 12 |
| TG | BBJ | EAS | 111667 | | 0.056 | 0.7 | T | 1.07 | 0.00 | 0.00 | 0.000 | 2 |
| | MVP | EUR | 24009 | 150983 | 0.01 | 0.7 | F | 1.32 | 0.25 | | 0.012 | 13 |
| | MVP | AFR | 5273 | 42485 | 1 | 1.4 | T | 1.07 | 0.05 | | 0.002 | 13 |
| | MVP | HISP | 1925 | 18285 | 0.0032 | 1.4 | T | 1.07 | 0.03 | | 0.002 | 13 |
| | FinnGen | FIN | 11924 | 288638 | 0.0056 | 0.7 | F | 1.16 | 0.08 | | 0.004 | 4 |
| PAD | BBJ | EAS | 4112 | 173601 | 0.0032 | 0.7 | F | 1.13 | 0.09 | 0.04 | 0.004 | 2 |
| | MEGA-STROKE | EUR | 40585 | 406111 | 0.0032 | 0.7 | T | 1.17 | 0.17 | | 0.030 | 14 |
| Stroke | BBJ | EAS | 22664 | 152022 | 0.56 | 1.4 | T | 1.06 | 0.05 | 0.07 | 0.010 | 2 |

| | | | | | | | | | | | | |
|---|---|---|---|---|---|---|---|---|---|---|---|---|
| | GBMI | AFR | 1161 | 24416 | 0.018 | 1.4 | T | 1.04 | 0.05 | | 0.010 | [15] |
| | MVP | EUR | 567460 | | 0.0001 | 0.7 | T | 1.05 | 0.04 | | 0.025 | [16] |
| | MVP | AFR | 13842 | | 0.018 | 1 | F | 1.04 | 0.03 | | 0.019 | [16] |
| CKD | BBJ | EAS | 150266 | | 0.0056 | 0.7 | T | 1.02 | 0.00 | 0.06 | 0.000 | [2] |

Traits: CAD – coronary artery disease; BMI – body mass index; DBP – diastolic blood pressure; SBP – systolic blood pressure; DM – diabetes mellitus; LDL-C – low density lipoprotein cholesterol; HDL-C – high-density lipoprotein cholesterol; TG – triglycerides; PAD – peripheral artery disease; CKD – chronic kidney disease.

Consortia: CARDIOGRAMplusC4D no UKBB – Coronary ARtery DIsease Genomewide Replication and Meta-analysis plus The Coronary Artery Disease Genetics consortium excluding UK Biobank; BBJ – Biobank Japan; GIANT – Genetic Investigation of ANthropometric Traits; MVP – Million Veteran Program; AGEN-T2D – Asian Genetic Epidemiology Network Type 2 Diabetes Consortium; GLGC – Global Lipids Genetics Consortium excluding the UK Biobank; GBMI – Global Biobank Meta-analysis Initiative

Ancestries: AFR – African; EAS – East Asian; EUR – European; FIN – Finnish; SAS – South Asian

LDPred2 parameters: ρ – tuning parameter to model the proportion of variants assumed to be causal; $h^2$ scale – factor by which heritability is scaled; is sparse LD – whether a sparse linkage disequilibrium matrix is applied, if true some variant effect estimates are fit to zero. A total of 102 combinations of these three parameters were possible and led to generation of 102 polygenic scores for each input ancestry-stratified set of GWAS summary statistics. The parameters leading to the best individually performing polygenic score in predicting CAD in a training dataset of 116,649 participants in the UK Biobank were chosen, specific to each ancestry-specific GWAS.

Mixing weights: Layer 1 – weight determined from a logistic regression model predicting CAD in a UK Biobank training dataset by combining different ancestry-specific GPS from GWAS for a specific trait. Layer 2 - weight determined from a logistic regression model predicting CAD in the same UKB training dataset by combining different trait-specific scores from the Layer 1 step. Final weight - overall mixing weight incorporating proportional weights from layers 1 and 2, normalized to 100%. The final $GPS_{Mult}$ score was a linear combination of all the input scores according to the final weight. The dataset for training the models and for estimating the mixing weight was from 116,649 participants in the UK Biobank. All the regression models for predicting CAD were adjusted for age, sex, genotyping array, and the first ten principal components of ancestry.

**Table 2:** Summary of $GPS_{Mult}$ training and validation datasets

| Stage | Training | Validation | | | | | | |
|---|---|---|---|---|---|---|---|---|
| Cohort | UK Biobank | UK Biobank | | | | MVP | | G&H |
| Ancestry | European | African | East Asian | European | South Asian | African | European | South Asian |
| N | 116649 | 7281 | 1464 | 308264 | 8982 | 33096 | 124467 | 16874 |
| Age (SD) | 57.5 (7.9) | 52.4 (8.0) | 53.0 (7.6) | 57.3 (8.0) | 53.8 (8.5) | 56.1 (12.4) | 60.8 (13.3) | 40.6 (14.5) |
| Male Sex (%) | 47.5% | 43.5% | 37.2% | 45.6% | 54.1% | 84.1% | 91.3% | 45.9% |
| Case | 4412 | 124 | 22 | 10492 | 542 | 4831 | 29171 | 853 |
| Control | 112237 | 7157 | 1442 | 297772 | 8440 | 28265 | 95296 | 16021 |

Baseline demographics of participants included in $GPS_{Mult}$ training and validation. These individuals were not included in genome-wide association data used to construct $GPS_{Mult}$, and individuals in training and validation of $GPS_{Mult}$ are distinct.

**Table 3:** Performance of polygenic scores for CAD from Polygenic Score Catalog in the UK Biobank Study

| Score Name | Identifier | Publication | Beta | SE | P | OR/SD [95% CI] | Population |
|---|---|---|---|---|---|---|---|
| GRS28 | PGS000200 | Tikkanen E et al. Arterioscler Thromb Vasc Biol (2013) | 0.27 | 0.29 | <0.001 | 1.31 [1.31-1.34] | EUR |
| GRS27 | PGS000010 | Mega JL et al. Lancet (2015) | 0.3 | 0.32 | <0.001 | 1.35 [1.35-1.38] | EUR |
| GRS50 | PGS000011 | Tada H et al. Eur Heart J (2015) | 0.29 | 0.31 | <0.001 | 1.34 [1.34-1.37] | EUR |
| GRS49K | PGS000012 | Abraham G et al. Eur Heart J (2016) | 0.36 | 0.39 | <0.001 | 1.44 [1.44-1.47] | EUR |
| CHD57 | PGS000057 | Natarajan P et al. Circulation (2017) | 0.29 | 0.31 | <0.001 | 1.34 [1.34-1.37] | EUR |
| GRS_CAD | PGS000019 | Paquette M et al. J Clin Lipidol (2017) | 0.4 | 0.42 | <0.001 | 1.49 [1.49-1.52] | EUR |
| $GPS_{2018}$ | PGS000013 | Khera AV et al. Nat Genet (2018) | 0.54 | 0.57 | <0.001 | 1.72 [1.72-1.76] | EUR |
| metaGRS_CAD | PGS000018 | Inouye M et al. J Am Coll Cardiol (2018) | 0.57 | 0.59 | <0.001 | 1.77 [1.77-1.8] | EUR |
| CHD46 | PGS000059 | Hajek C et al. Circ Genom Precis Med (2018) | 0.24 | 0.26 | <0.001 | 1.27 [1.27-1.3] | EUR |
| CAD_EJ2020 | PGS000116 | Elliott J et al. JAMA (2020) | 0.42 | 0.44 | <0.001 | 1.53 [1.53-1.56] | EUR |
| PRS_COMBINED | PGS000749 | Gola D et al. Circ Genom Precis Med (2020) | 0.4 | 0.42 | <0.001 | 1.49 [1.49-1.52] | EUR |
| PRS_CHD | PGS000329 | Mars N et al. Nat Med (2020) | 0.5 | 0.52 | <0.001 | 1.64 [1.64-1.68] | EUR |
| GPS_CAD_SA | PGS000296 | Wang M et al. J Am Coll Cardiol (2020) | 0.51 | 0.54 | <0.001 | 1.67 [1.67-1.71] | EUR |
| PRS70_CAD | PGS000349 | Pechlivanis S et al. BMC Med Genet (2020) | 0.14 | 0.16 | <0.001 | 1.14 [1.14-1.17] | EUR |
| GRS_Metabo | PGS000818 | Bauer A et al. Genet Epidemiol (2021) | 0.39 | 0.41 | <0.001 | 1.48 [1.48-1.51] | EUR |
| $AnnoPred_{CAD}$ | PGS001355 | Ye Y et al. Circ Genom Precis Med (2021) | 0.57 | 0.59 | <0.001 | 1.76 [1.76-1.8] | EUR |
| metaPRS_CAD | PGS002262 | Lu X et al. Eur Heart J (2022) | 0.26 | 0.28 | <0.001 | 1.30 [1.3-1.32] | EUR |
| CHD_PRSCS | PGS001780 | Tamlander M et al. Commun Biol (2022) | 0.55 | 0.57 | <0.001 | 1.73 [1.73-1.77] | EUR |
| $GPS_{Mult}$ | - | - | 0.75 | 0.77 | <0.001 | 2.12 [2.12-2.17] | EUR |

A series of prior published scores or CAD from the Polygenic Score Catalog that did not include any UK Biobank participants in the GWAS data for score development were compared with $GPS_{Mult}$ within the validation cohort of European ancestry individuals from the UK Biobank. The odds ratio for prevalent coronary artery disease (CAD) risk per standard deviation of the polygenic score and 95% confidence interval were assessed in a logistic regression model adjusted for age, sex, genotyping array, and the first ten principal components of ancestry.

**Table 4:** Baseline risk factor distributions and correlations with $GPS_{Mult}$

| | Prevalent CAD | Incident CAD | No CAD | All | Correlation |
|---|---|---|---|---|---|
| **n** | 11180 | 11898 | 302913 | 325991 | |
| **LDL Cholesterol* (mg/dl, mean (SD))** | 143.78 (35.76) | 156.08 (35.81) | 145.25 (32.95) | 145.60 (33.22) | 0.152 |
| **HDL Cholesterol* (mg/dl, mean (SD))** | 46.20 (12.16) | 50.49 (13.19) | 56.72 (14.81) | 56.13 (14.84) | -0.115 |
| **Triglycerides* (mg/dl, median [IQR])** | 170.17 [118.87, 243.84] | 167.25 [117.68, 241.03] | 131.98 [92.65, 192.47] | 134.37 [93.98, 196.19] | 0.108 |
| **Systolic Blood Pressure (mm Hg, mean (SD))** | 140.00 (19.86) | 147.28 (20.06) | 139.37 (19.56) | 139.68 (19.64) | 0.081 |
| **Diastolic Blood Pressure (mm Hg, mean (SD))** | 78.94 (10.93) | 84.57 (10.94) | 82.27 (10.66) | 82.24 (10.71) | 0.071 |
| **Body-mass Index ($kg/m^2$, mean (SD))** | 29.13 (4.77) | 28.58 (4.79) | 27.26 (4.74) | 27.37 (4.76) | 0.108 |
| **Diabetes Mellitus# (%)** | 2197 (19.7) | 1460 (12.3) | 14012 (4.6) | 17669 (5.4) | 0.028 |
| **Chronic Kidney Disease# (%)** | 1621 (15.2) | 1134 (10.0) | 11484 (4.0) | 14239 (4.6) | 0.091 |

The baseline variable distributions for traditional CAD risk factors stratified by CAD case status. Correlations between the $GPS_{Mult}$ and risk factors across the entire validation population, adjusted for top 10 principal components and traditional risk factors were assessed using Pearson's correlation for continuous traits and #point biserial correlation for binary traits. The P value for each association was less than $2 \times 10^{-12}$. *LDL-C, HDL-C, and triglyceride values were adjusted for cholesterol-lowering medication status, as previously described.[17] Diagnosis of diabetes was based on self-report, primary care records or hospitalization records confirming a clinical diagnosis, self-reported consumption of medications to treat diabetes, or glycohemoglobin >6.5% at enrollment and CKD was defined as eGFRcys <60 mL/min, as previously described.[18] *Adjusted values if individual taking cholesterol lowering medications as described in Patel et al. *JAMA Network Open* 2020. For example, in the case of statin intake, lower density cholesterol was divided by 0.7 and triglycerides by 0.85.

**Table 5:** Baseline characteristics according to high $GPS_{Mult}$

| | Bottom 80% $GPS_{Mult}$ | Top 20% $GPS_{Mult}$ | p |
|---|---|---|---|
| Number of individuals | 224,292 | 56,424 | |
| Prevalent coronary artery disease (%) | 5,496 (2.5) | 4,155 (7.4) | <0.001 |
| Age, years (mean, SD) | 57.5 (7.9) | 57.2 (9.0) | <0.001 |
| Male sex (%) | 102,639 (45.8) | 25,926 (45.9) | 0.428 |
| Hypertension (%) | 58,635 (27.2) | 20,305 (37.9) | <0.001 |
| Diabetes Mellitus (%) | 9,219 (4.1) | 4,653 (8.2) | <0.001 |
| *LDL cholesterol, mg/dL (mean (SD)) | 137.7 (33.0) | 140.5 (35.9) | <0.001 |
| *HDL cholesterol, mg/dL (mean (SD)) | 57.0 (14.9) | 53.9 (14.3) | <0.001 |
| *Triglycerides, mg/dL (median [IQR]) | 129.4 [91.6, 186.8] | 142.3 [100.7, 204.9] | <0.001 |
| Current smoking (%) | 20,365 (9.1) | 5,866 (10.4) | <0.001 |
| Family history of heart disease (%) | 95,049 (42.4) | 29,936 (53.1) | <0.001 |
| Body mass index, kg/m$^2$ (mean (SD)) | 27.2 (4.7) | 28.1 (5.0) | <0.001 |
| Lipid-lowering therapy (%) | 31,201 (13.9) | 13,892 (24.6) | <0.001 |

Baseline characteristics according to high $GPS_{Mult}$ status, defined as the top 20% of the distribution empirically shown to be at ≥3-fold risk of coronary artery disease. LDL: low-density lipoprotein; HDL: high-density lipoprotein. *LDL-C, HDL-C, and triglyceride values were adjusted for cholesterol-lowering medication status, as previously described.[17]

**Table 6:** Performance of polygenic scores for CAD from Polygenic Score Catalog in the Million Veteran Program Study

| Score Name | Identifier | Publication | Beta | SE | P | OR/SD [95% CI] | Population |
|---|---|---|---|---|---|---|---|
| GRS28 | PGS000200 | Tikkanen E et al. Arterioscler Thromb Vasc Biol (2013) | 0.053 | 0.016 | 0.001 | 1.05 [1.02-1.09] | AFR |
| GRS27 | PGS000010 | Mega JL et al. Lancet (2015) | 0.066 | 0.016 | <0.001 | 1.07 [1.04-1.1] | AFR |
| GRS50 | PGS000011 | Tada H et al. Eur Heart J (2015) | 0.073 | 0.016 | <0.001 | 1.08 [1.04-1.11] | AFR |
| GRS49K | PGS000012 | Abraham G et al. Eur Heart J (2016) | 0.082 | 0.016 | <0.001 | 1.09 [1.05-1.12] | AFR |
| CHD57 | PGS000057 | Natarajan P et al. Circulation (2017) | 0.066 | 0.016 | <0.001 | 1.07 [1.03-1.1] | AFR |
| GRS_CAD | PGS000019 | Paquette M et al. J Clin Lipidol (2017) | 0.115 | 0.016 | <0.001 | 1.12 [1.09-1.16] | AFR |
| $GPS_{2018}$ | PGS000013 | Khera AV et al. Nat Genet (2018) | 0.128 | 0.016 | <0.001 | 1.14 [1.1-1.17] | AFR |
| CAD_GRS_204 | PGS000058 | Morieri ML et al. Diabetes Care (2018) | 0.117 | 0.016 | <0.001 | 1.12 [1.09-1.16] | AFR |
| metaGRS_CAD | PGS000018 | Inouye M et al. J Am Coll Cardiol (2018) | 0.155 | 0.016 | <0.001 | 1.17 [1.13-1.21] | AFR |
| CHD46 | PGS000059 | Hajek C et al. Circ Genom Precis Med (2018) | 0.060 | 0.016 | <0.001 | 1.06 [1.03-1.1] | AFR |
| 157SNP_GRS | PGS000798 | Severance LM et al. J Cardiovasc Comput Tomogr (2019) | 0.086 | 0.016 | <0.001 | 1.09 [1.06-1.13] | AFR |
| CAD_EJ2020 | PGS000116 | Elliott J et al. JAMA (2020) | 0.114 | 0.016 | <0.001 | 1.12 [1.08-1.16] | AFR |
| PRS_DE | PGS000748 | Gola D et al. Circ Genom Precis Med (2020) | 0.118 | 0.016 | <0.001 | 1.13 [1.09-1.16] | AFR |
| PRS_COMBINED | PGS000749 | Gola D et al. Circ Genom Precis Med (2020) | 0.112 | 0.016 | <0.001 | 1.12 [1.08-1.16] | AFR |
| PRS_CHD | PGS000329 | Mars N et al. Nat Med (2020) | 0.144 | 0.016 | <0.001 | 1.15 [1.12-1.19] | AFR |
| GPS_CAD_SA | PGS000296 | Wang M et al. J Am Coll Cardiol (2020) | 0.158 | 0.016 | <0.001 | 1.17 [1.13-1.21] | AFR |
| PRS70_CAD | PGS000349 | Pechlivanis S et al. BMC Med Genet (2020) | 0.028 | 0.016 | 0.0812 | 1.03 [1-1.06] | AFR |
| $MetaPRS_{CAD}$ | PGS000337 | Koyama S et al. Nat Genet (2020) | 0.167 | 0.016 | <0.001 | 1.18 [1.14-1.22] | AFR |
| PRS176_CHD | PGS000899 | Feitosa MF et al. Circ Genom Precis Med (2021) | 0.109 | 0.016 | <0.001 | 1.11 [1.08-1.15] | AFR |
| GRS_Metabo | PGS000818 | Bauer A et al. Genet Epidemiol (2021) | 0.101 | 0.016 | <0.001 | 1.11 [1.07-1.14] | AFR |

| | | | | | | | |
|---|---|---|---|---|---|---|---|
| AnnoPred$_{CAD}$ | PGS001355 | Ye Y et al. Circ Genom Precis Med (2021) | 0.161 | 0.016 | <0.001 | 1.18 [1.14-1.21] | AFR |
| portability-ldpred2 | PGS002048 | Prive F et al. Am J Hum Genet (2022) | 0.129 | 0.016 | <0.001 | 1.14 [1.1-1.17] | AFR |
| metaPRS_CAD | PGS002262 | Lu X et al. Eur Heart J (2022) | 0.061 | 0.016 | <0.001 | 1.06 [1.03-1.1] | AFR |
| PRSCS$_{CHD}$ | PGS001780 | Tamlander M et al. Commun Biol (2022) | 0.166 | 0.016 | <0.001 | 1.18 [1.14-1.22] | AFR |
| GBE_HC942 | PGS000962 | Tanigawa Y et al. PLoS Genet (2022) | 0.088 | 0.016 | <0.001 | 1.09 [1.06-1.13] | AFR |
| ldpred_cad | PGS002244 | Mars N et al. Cell Genom (2022) | 0.139 | 0.016 | <0.001 | 1.15 [1.11-1.19] | AFR |
| PRS_2022 | PGS003356 | Aragam KA et al. Nat Genet (2022) | 0.160 | 0.016 | <0.001 | 1.17 [1.14-1.21] | AFR |
| GPSMult | - | - | 0.222 | 0.016 | <0.001 | 1.25 [1.21-1.29] | AFR |
| GRS28 | PGS000200 | Tikkanen E et al. Arterioscler Thromb Vasc Biol (2013) | 0.185 | 0.007 | <0.001 | 1.2 [1.19-1.22] | EUR |
| GRS27 | PGS000010 | Mega JL et al. Lancet (2015) | 0.214 | 0.007 | <0.001 | 1.24 [1.22-1.26] | EUR |
| GRS50 | PGS000011 | Tada H et al. Eur Heart J (2015) | 0.195 | 0.007 | <0.001 | 1.21 [1.2-1.23] | EUR |
| GRS49K | PGS000012 | Abraham G et al. Eur Heart J (2016) | 0.249 | 0.007 | <0.001 | 1.28 [1.27-1.3] | EUR |
| CHD57 | PGS000057 | Natarajan P et al. Circulation (2017) | 0.118 | 0.007 | <0.001 | 1.13 [1.11-1.14] | EUR |
| GRS_CAD | PGS000019 | Paquette M et al. J Clin Lipidol (2017) | 0.273 | 0.007 | <0.001 | 1.31 [1.3-1.33] | EUR |
| GPS_CAD | PGS000013 | Khera AV et al. Nat Genet (2018) | 0.370 | 0.007 | <0.001 | 1.45 [1.43-1.47] | EUR |
| CAD_GRS_204 | PGS000058 | Morieri ML et al. Diabetes Care (2018) | 0.326 | 0.007 | <0.001 | 1.39 [1.37-1.41] | EUR |
| metaGRS_CAD | PGS000018 | Inouye M et al. J Am Coll Cardiol (2018) | 0.385 | 0.007 | <0.001 | 1.47 [1.45-1.49] | EUR |
| CHD46 | PGS000059 | Hajek C et al. Circ Genom Precis Med (2018) | 0.165 | 0.007 | <0.001 | 1.18 [1.16-1.2] | EUR |
| 157SNP_GRS | PGS000798 | Severance LM et al. J Cardiovasc Comput Tomogr (2019) | 0.289 | 0.007 | <0.001 | 1.33 [1.32-1.35] | EUR |
| CAD_EJ2020 | PGS000116 | Elliott J et al. JAMA (2020) | 0.295 | 0.007 | <0.001 | 1.34 [1.32-1.36] | EUR |
| PRS_DE | PGS000748 | Gola D et al. Circ Genom Precis Med (2020) | 0.273 | 0.007 | <0.001 | 1.31 [1.3-1.33] | EUR |
| PRS_COMBINED | PGS000749 | Gola D et al. Circ Genom Precis Med (2020) | 0.275 | 0.007 | <0.001 | 1.32 [1.3-1.33] | EUR |

| | | | | | | | |
|---|---|---|---|---|---|---|---|
| PRS_CHD | PGS000329 | Mars N et al. Nat Med (2020) | 0.367 | 0.007 | <0.001 | 1.44 [1.42-1.46] | EUR |
| GPS_CAD_SA | PGS000296 | Wang M et al. J Am Coll Cardiol (2020) | 0.356 | 0.007 | <0.001 | 1.43 [1.41-1.45] | EUR |
| PRS70_CAD | PGS000349 | Pechlivanis S et al. BMC Med Genet (2020) | 0.104 | 0.007 | <0.001 | 1.11 [1.09-1.13] | EUR |
| MetaPRS$_{CAD}$ | PGS000337 | Koyama S et al. Nat Genet (2020) | 0.418 | 0.007 | <0.001 | 1.52 [1.5-1.54] | EUR |
| PRS176_CHD | PGS000899 | Feitosa MF et al. Circ Genom Precis Med (2021) | 0.327 | 0.007 | <0.001 | 1.39 [1.37-1.41] | EUR |
| GRS_Metabo | PGS000818 | Bauer A et al. Genet Epidemiol (2021) | 0.258 | 0.007 | <0.001 | 1.29 [1.28-1.31] | EUR |
| AnnoPred$_{CAD}$ | PGS001355 | Ye Y et al. Circ Genom Precis Med (2021) | 0.383 | 0.007 | <0.001 | 1.47 [1.45-1.49] | EUR |
| portability-ldpred2 | PGS002048 | Prive F et al. Am J Hum Genet (2022) | 0.379 | 0.007 | <0.001 | 1.46 [1.44-1.48] | EUR |
| metaPRS_CAD | PGS002262 | Lu X et al. Eur Heart J (2022) | 0.207 | 0.007 | <0.001 | 1.23 [1.21-1.25] | EUR |
| PRSCS$_{CHD}$ | PGS001780 | Tamlander M et al. Commun Biol (2022) | 0.372 | 0.007 | <0.001 | 1.45 [1.43-1.47] | EUR |
| GBE_HC942 | PGS000962 | Tanigawa Y et al. PLoS Genet (2022) | 0.287 | 0.007 | <0.001 | 1.33 [1.31-1.35] | EUR |
| ldpred_cad | PGS002244 | Mars N et al. Cell Genom (2022) | 0.344 | 0.007 | <0.001 | 1.41 [1.39-1.43] | EUR |
| PRS$_{2022}$ | PGS003356 | Aragam KA et al. Nat Genet (2022) | 0.477 | 0.008 | <0.001 | 1.61 [1.59-1.64] | EUR |
| GPS$_{Mult}$ | - | - | 0.542 | 0.008 | <0.001 | 1.72 [1.69-1.75] | EUR |

The odds ratio for prevalent coronary artery disease (CAD) risk per standard deviation of the polygenic score and 95% confidence interval were validated in a logistic regression model adjusted for age, sex, genotyping array, and the first ten principal components of ancestry in a holdout cohort of individuals of the Million Veteran Project using prior published scores from the Polygenic Score Catalog. AFR: African ancestry; EUR: European ancestry.

**Table 7:** Performance of polygenic scores for CAD from Polygenic Score Catalog in the Genes & Health Study

| Score Name | Identifier | Publication | Beta | SE | P | OR/SD [95% CI] | Population |
|---|---|---|---|---|---|---|---|
| GRS28 | PGS000200 | Tikkanen E et al. Arterioscler Thromb Vasc Biol (2013) | 0.169 | 0.040 | <0.001 | 1.18 [1.09-1.28] | SAS |
| GRS27 | PGS000010 | Mega JL et al. Lancet (2015) | 0.159 | 0.040 | <0.001 | 1.17 [1.08-1.27] | SAS |
| GRS50 | PGS000011 | Tada H et al. Eur Heart J (2015) | 0.108 | 0.039 | 0.006 | 1.11 [1.03-1.2] | SAS |
| GRS49K | PGS000012 | Abraham G et al. Eur Heart J (2016) | 0.288 | 0.044 | <0.001 | 1.33 [1.22-1.45] | SAS |
| CHD57 | PGS000057 | Natarajan P et al. Circulation (2017) | 0.105 | 0.040 | 0.009 | 1.11 [1.03-1.2] | SAS |
| GRS_CAD | PGS000019 | Paquette M et al. J Clin Lipidol (2017) | 0.218 | 0.040 | <0.001 | 1.24 [1.15-1.34] | SAS |
| $GPS_{2018}$ | PGS000013 | Khera AV et al. Nat Genet (2018) | 0.285 | 0.042 | <0.001 | 1.33 [1.23-1.44] | SAS |
| CAD_GRS_204 | PGS000058 | Morieri ML et al. Diabetes Care (2018) | 0.229 | 0.040 | <0.001 | 1.26 [1.16-1.36] | SAS |
| metaGRS_CAD | PGS000018 | Inouye M et al. J Am Coll Cardiol (2018) | 0.327 | 0.044 | <0.001 | 1.39 [1.27-1.51] | SAS |
| CHD46 | PGS000059 | Hajek C et al. Circ Genom Precis Med (2018) | 0.111 | 0.040 | 0.006 | 1.12 [1.03-1.21] | SAS |
| 157SNP_GRS | PGS000798 | Severance LM et al. J Cardiovasc Comput Tomogr (2019) | 0.244 | 0.041 | <0.001 | 1.28 [1.18-1.38] | SAS |
| CAD_EJ2020 | PGS000116 | Elliott J et al. JAMA (2020) | 0.327 | 0.042 | <0.001 | 1.39 [1.28-1.5] | SAS |
| PRS_COMBINED | PGS000749 | Gola D et al. Circ Genom Precis Med (2020) | 0.328 | 0.041 | <0.001 | 1.39 [1.28-1.51] | SAS |
| PRS_DE | PGS000748 | Gola D et al. Circ Genom Precis Med (2020) | 0.306 | 0.041 | <0.001 | 1.36 [1.25-1.47] | SAS |
| PRS_CHD | PGS000329 | Mars N et al. Nat Med (2020) | 0.310 | 0.041 | <0.001 | 1.36 [1.26-1.48] | SAS |
| GPS_CAD_SA | PGS000296 | Wang M et al. J Am Coll Cardiol (2020) | 0.338 | 0.041 | <0.001 | 1.4 [1.29-1.52] | SAS |
| PRS70_CAD | PGS000349 | Pechlivanis S et al. BMC Med Genet (2020) | 0.072 | 0.040 | 0.069 | 1.08 [0.99-1.16] | SAS |
| $MetaPRS_{CAD}$ | PGS000337 | Koyama S et al. Nat Genet (2020) | 0.428 | 0.042 | <0.001 | 1.53 [1.41-1.67] | SAS |
| PRS176_CHD | PGS000899 | Feitosa MF et al. Circ Genom Precis Med (2021) | 0.251 | 0.041 | <0.001 | 1.29 [1.19-1.39] | SAS |
| GRS_Metabo | PGS000818 | Bauer A et al. Genet Epidemiol (2021) | 0.209 | 0.040 | <0.001 | 1.23 [1.14-1.33] | SAS |

| | | | | | | | |
|---|---|---|---|---|---|---|---|
| $AnnoPred_{CAD}$ | PGS001355 | Ye Y et al. Circ Genom Precis Med (2021) | 0.352 | 0.041 | <0.001 | 1.42 [1.31-1.54] | SAS |
| portability-ldpred2 | PGS002048 | Prive F et al. Am J Hum Genet (2022) | 0.380 | 0.041 | <0.001 | 1.46 [1.35-1.59] | SAS |
| metaPRS_CAD | PGS002262 | Lu X et al. Eur Heart J (2022) | 0.193 | 0.041 | <0.001 | 1.21 [1.12-1.31] | SAS |
| $PRSCS_{CHD}$ | PGS001780 | Tamlander M et al. Commun Biol (2022) | 0.420 | 0.042 | <0.001 | 1.52 [1.4-1.65] | SAS |
| GBE_HC942 | PGS000962 | Tanigawa Y et al. PLoS Genet (2022) | 0.180 | 0.040 | <0.001 | 1.2 [1.11-1.29] | SAS |
| ldpred_cad | PGS002244 | Mars N et al. Cell Genom (2022) | 0.301 | 0.042 | <0.001 | 1.35 [1.25-1.47] | SAS |
| $PRS_{2022}$ | PGS003356 | Aragam KA et al. Nat Genet (2022) | 0.469 | 0.042 | <0.001 | 1.6 [1.47-1.73] | SAS |
| $GPS_{Mult}$ | - | - | 0.606 | 0.042 | <0.001 | 1.83 [1.69-1.99] | SAS |

The odds ratio for prevalent coronary artery disease (CAD) risk per standard deviation of the polygenic score and 95% confidence interval were validated in a logistic regression model adjusted for age, sex, genotyping array, and the first ten principal components of ancestry in a holdout cohort of individuals of the Genes & Health study using prior published scores from the Polygenic Score Catalog. SAS: South Asian ancestry.

**Table 8:** Model C-statistics by ancestry in the UK Biobank Study

| Ancestry | All | African | East Asian | European | South Asian |
|---|---|---|---|---|---|
| **Age+Sex** | 0.71 (0.706-0.715) | 0.708 (0.666-0.75) | 0.775 (0.694-0.856) | 0.708 (0.703-0.712) | 0.712 (0.692-0.732) |
| **PCE** | 0.739 (0.735-0.744) | 0.749 (0.707-0.791) | 0.803 (0.71-0.895) | 0.739 (0.734-0.743) | 0.739 (0.717-0.76) |
| **$PCE+GPS_{Mult}+PCE*GPS_{Mult}$** | 0.763 (0.759-0.768) | 0.753 (0.712-0.794) | 0.882 (0.843-0.921) | 0.762 (0.758-0.767) | 0.764 (0.743-0.784) |
| **QRISK3** | 0.746 (0.741-0.75) | 0.747 (0.698-0.796) | 0.811 (0.73-0.893) | 0.744 (0.74-0.749) | 0.745 (0.722-0.769) |
| **$QRISK3+GPS_{Mult}+QRISK3*GPS_{Mult}$** | 0.769 (0.764-0.773) | 0.759 (0.712-0.806) | 0.882 (0.834-0.929) | 0.768 (0.763-0.772) | 0.777 (0.756-0.799) |

C-statistics stratified by ancestry are based on 10-year follow-up events from Cox regression models of listed variables, the first ten principal components of genetic ancestry, and genotyping array. ACC/AHA Pooled Cohort Equations (PCE) and QISK3 include age and sex variables in their risk estimation.

**Table 9:** Model net reclassification by ancestry in the UK Biobank Study

| NRI | | Status | All | African | East Asian | European | South Asian |
|---|---|---|---|---|---|---|---|
| Categorical | PCE | Full | 0.070 (0.059-0.082) | 0.010 (-0.016-0.035) | 0.000 (-0.008-0.176) | 0.073 (0.063-0.081) | 0.067 (0.019-0.147) |
| | | Cases | 0.081 (0.069-0.094) | 0.010 (-0.016-0.034) | 0.000 (0.000-0.179) | 0.083 (0.074-0.092) | 0.102 (0.043-0.188) |
| | | Noncases | -0.011 (-0.012 - -0.009) | 0.00 (-0.003-0.002) | 0000 (-0.008-0.003) | -0.011 (-0.012-0.009) | -0.035 (-0.053-0.018) |
| | QRISK3 | Full | 0.034 (0.028-0.040) | 0.011 (-0.014-0.058) | -0.001 (-0.003-0.002) | 0.036 (0.029-0.044) | 0.025 (-0.011-0.101) |
| | | Cases | 0.038 (0.032-0.045) | 0.012 (-0.012-0.059) | 0.000 (0.000-0.000) | 0.040 (0.033-0.049) | 0.035 (-0.004-0.124) |
| | | Noncases | -0.004 (-0.005 - -0.003) | -0.001 (-0.003-0.001) | -0.001 (-0.003-0.002) | -0.005 (-0.006-0.004) | -0.011 (-0.023-0.003) |
| Continuous | PCE | Full | 0.405 (0.379-0.424) | 0.057 (-0.211-0.291) | 0.427 (-0.249-0.849) | 0.412 (0.388-0.431) | 0.366 (0.269-0.463) |
| | | Cases | 0.200 (0.182-0.215) | -0.026 (-0.245-0.184) | 0.285 (-0.402-0.599) | 0.206 (0.186-0.221) | 0.175 (0.104-0.257) |
| | | Noncases | 0.205 (0.197-0.213) | 0.084 (-0.012-0.183) | 0.142 (-0.007-0.347) | 0.207 (0.195-0.217) | 0.191 (0.158-0.233) |
| | QRISK3 | Full | 0.395 (0.373-0.418) | 0.173 (-0.207-0.673) | 0.300 (-0.338-0.780) | 0.403 (0.377-0.426) | 0.355 (0.232-0.456) |
| | | Cases | 0.195 (0.177-0.214) | 0.059 (-0.165-0.443) | 0.164 (-0.496-0.594) | 0.201 (0.183-0.218) | 0.173 (0.073-0.246) |
| | | Noncases | 0.200 (0.192-0.210) | 0.114 (-0.013-0.406) | 0.137 (-0.015-0.383) | 0.202 (0.193-0.210) | 0.183 (0.138-0.243) |

The improvement in the predictive performance of the addition of the $GPS_{Mult}$ to the ACC/AHA Pooled Cohort Equations (PCE) or QRISK3 was evaluated in each ancestry using continuous and categorised net reclassification improvement (NRI), with a risk probabilities threshold of 7.5% obtained with Kaplan-Meier estimates for a period of 10 years and confidence intervals (95%) obtained from 100-fold bootstrapping.

**Figure 1:** Sequential improvements in $R^2$ with $GPS_{Mult}$ in the UK Biobank Study

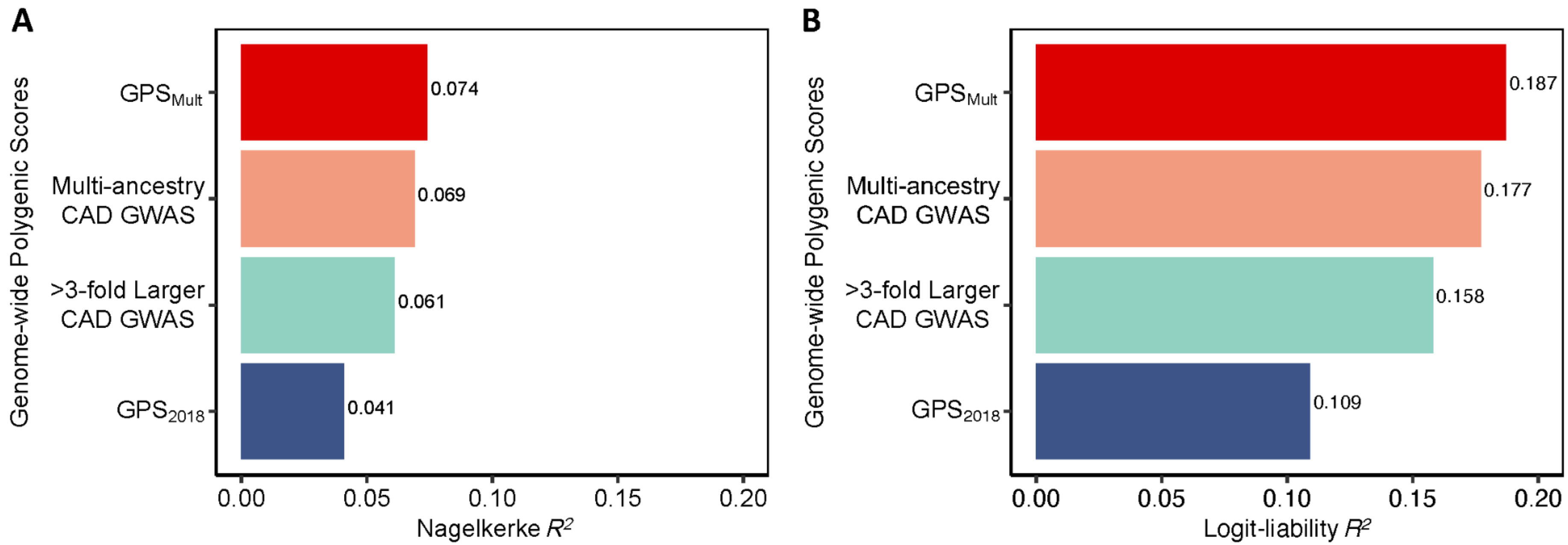

The proportion of phenotypic variance explained by the polygenic score predicting coronary artery disease (CAD) was calculated in the UK Biobank European ancestry validation cohort for each GPS score using the A: Nagelkerke's pseudo-$R^2$ metric, as the difference of the full model inclusive of the polygenic score plus age, sex, genotyping array, and the first ten principal components of ancestry minus $R^2$ for the covariates alone; and B: logit-liability $R^2$ metric, as described elsewhere.[19] $GPS_{2018}$ denotes previously published polygenic score for CAD.[20] >3-fold larger CAD GWAS designates metrics for polygenic score generated using summary statistics from the most recent Coronary ARtery DIsease Genome wide Replication and Meta-analysis plus The Coronary Artery Disease Genetics consortium analysis (CARDIOGRAMplusC4D) excluding the UKBB. Multi-ancestry CAD GWAS refers to the polygenic score generated by combining ancestry-specific polygenic scores generated using discovery data from Genes & Health, Biobank Japan, Million Veteran Program, FinnGen, and CARDIOGRAMplusC4D (excluding UKBB). $GPS_{Mult}$ designates polygenic score for CAD designed with summary statistics from multiple ancestries and multiple CAD-related traits.

**Figure 2:** $GPS_{Mult}$ performance by sex and age subgroups

| Subgroup | N CAD (%) | OR/SD | 95%−CI | P−Value |
|---|---|---|---|---|
| Men | 8,449 / 140,471 (6 %) | 2.20 | [2.15; 2.26] | <0.0001 |
| Women | 2,043 / 167,793 (1.2 %) | 1.94 | [1.86; 2.03] | <0.0001 |
| Ages 45−54 | 1,064 / 85,790 (1.2 %) | 2.17 | [2.04; 2.31] | <0.0001 |
| Ages 55−64 | 4,955 / 133,302 (3.7 %) | 2.18 | [2.11; 2.25] | <0.0001 |
| Ages 65−75 | 4,322 / 59,876 (7.2 %) | 2.08 | [2.01; 2.15] | <0.0001 |

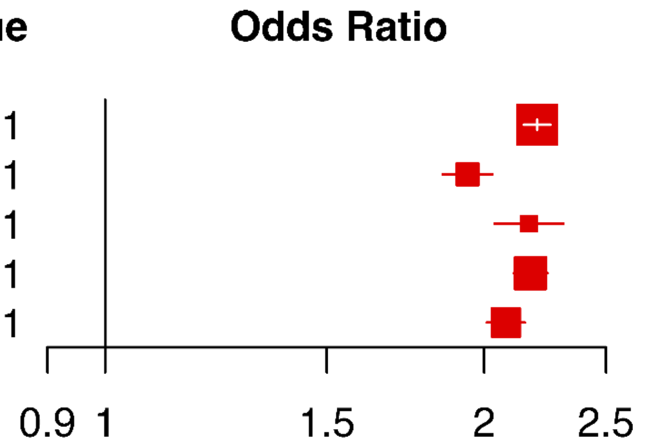

The odds ratio for prevalent coronary artery disease (CAD) risk per standard deviation of the $GPS_{Mult}$ was assessed in a logistic regression model adjusted for age, sex, genotyping array, and the first ten principal components of genetic ancestry in the European ancestry validation dataset of the UK biobank stratified by sex and age subgroups.

**Figure 3:** Equivalents of increased risk for incident coronary artery disease event identified by high $GPS_{Mult}$ in the UK Biobank Study

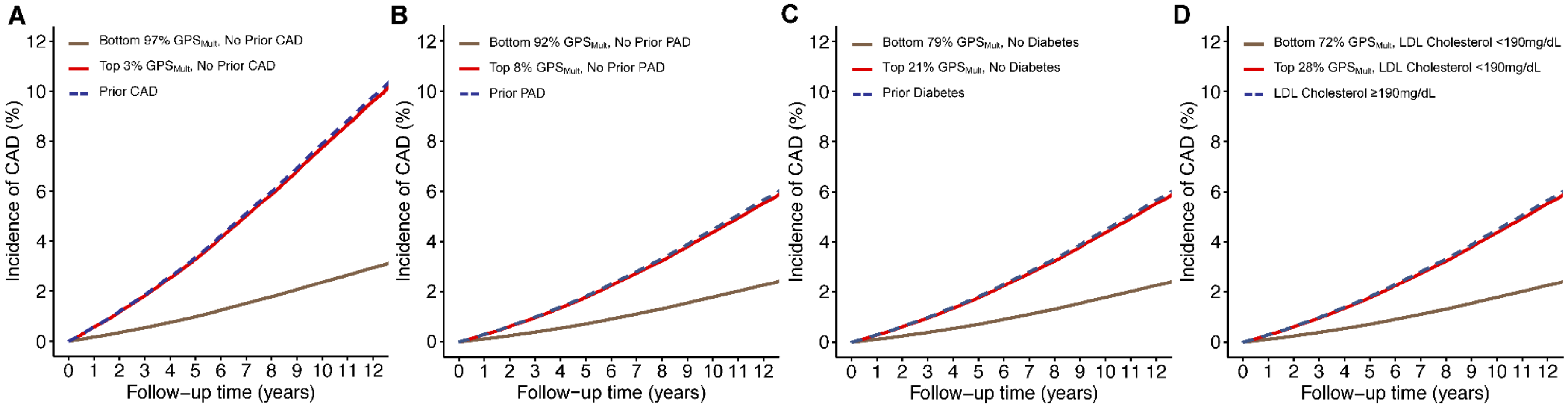

A: Cumulative incidence of coronary artery disease (CAD) events over length of follow-up stratified by presence of prior CAD, no prior CAD with $GPS_{Mult}$ in the bottom 97% or top 3% of the population, estimated using Cox proportional-hazards regression model adjusted for age, sex, genotyping array, and the first ten principal components of ancestry in the European ancestry UK Biobank validation dataset. B: Cumulative incidence of coronary artery disease (CAD) events over length of follow-up stratified by presence of prior peripheral artery disease (PAD) or no prior PAD with $GPS_{Mult}$ in the bottom 92% or top 8% of the population, estimated using Cox proportional-hazards regression model adjusted for age, sex, genotyping array, and the first ten principal components of ancestry in the European ancestry UK Biobank validation dataset. C: Cumulative incidence of CAD events over length of follow-up stratified by presence of prior diabetes mellitus (DM) or no prior DM with $GPS_{Mult}$ in the bottom 79% or top 21% of the population, estimated using Cox proportional-hazards regression model adjusted for age, sex, genotyping array, and the first ten principal components of ancestry in the European ancestry UK Biobank validation dataset. D: Cumulative incidence of CAD events over length of follow-up stratified by presence of prior severe hypercholesterolemia (estimated untreated low-density lipoprotein cholesterol, LDL-C 190 mg/dL or higher), or no prior hypercholesterolemia with $GPS_{Mult}$ in the bottom 72% or top 28% of the population, estimated using Cox proportional-hazards regression model adjusted for age, sex, genotyping array, and the first ten principal components of ancestry in the European ancestry UK Biobank validation dataset.

**Figure 4:** Decrease in risk for incident coronary artery disease identified by low $GPS_{Mult}$

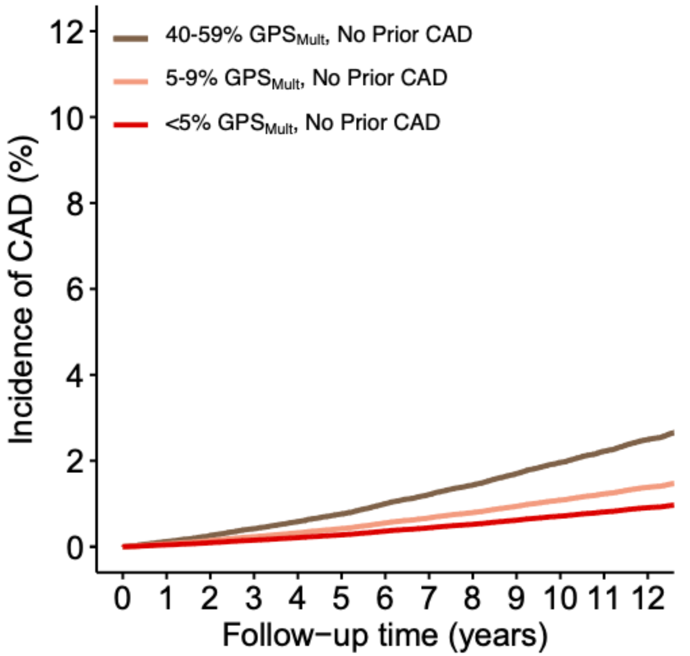

Cumulative CAD risk stratified by $GPS_{Mult}$ in the bottom 5%, 5-9%, and 40-59% of the population, estimated using Cox proportional-hazards regression model adjusted for age, sex, genotyping array, and the first ten principal components of ancestry in the European ancestry validation dataset of the UK Biobank Study. GPS: Genome-wide polygenic score.

**Figure 5:** Net reclassification for $GPS_{Mult}$ and CAD risk enhancers over PCE 10-year risk estimates

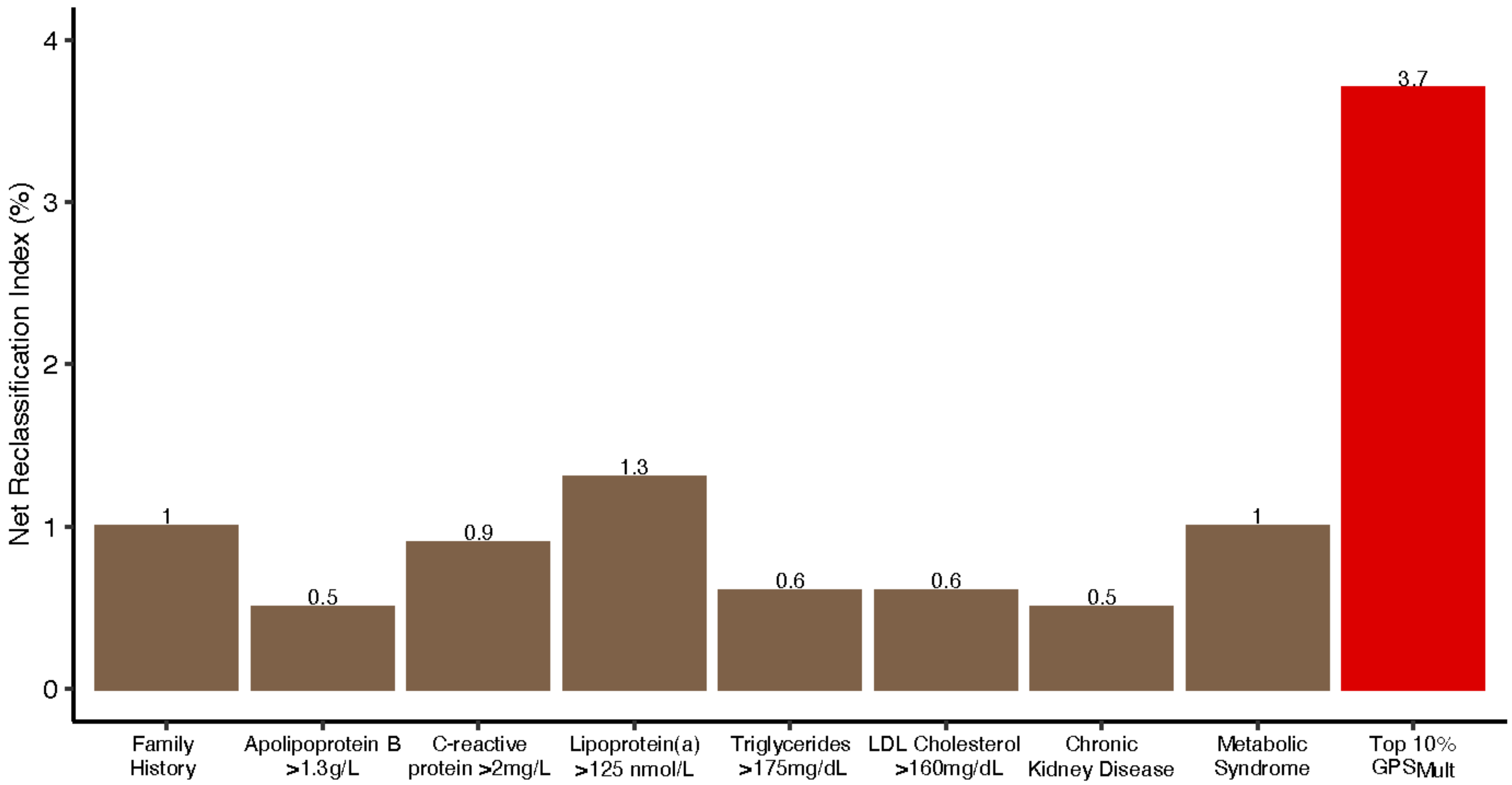

Net reclassification of coronary artery disease (CAD) cases and non-cases achieved by established CAD risk enhancing factors and $GPS_{Mult}$ when added to a baseline model of just the American Heart Association/American College of Cardiology Pooled Cohort Equations[21] in the European ancestry validation dataset of the UK Biobank.

**Figure 6:** Association of $GPS_{Mult}$ and risk factors with incident and recurrent coronary artery disease events

| Predictor | Event | HR/SD | 95%–CI | P–Value |
|---|---|---|---|---|
| $GPS_{Mult}$ | Incident | 1.73 | [1.70; 1.76] | <0.0001 |
| | Recurrent | 1.13 | [1.08; 1.18] | <0.0001 |
| Systolic Blood Pressure | Incident | 1.21 | [1.18; 1.23] | <0.0001 |
| | Recurrent | 1.20 | [1.15; 1.25] | <0.0001 |
| Diastolic Blood Pressure | Incident | 1.13 | [1.11; 1.15] | <0.0001 |
| | Recurrent | 1.11 | [1.06; 1.16] | <0.0001 |
| Glycated Hemoglobin | Incident | 1.20 | [1.17; 1.22] | <0.0001 |
| | Recurrent | 1.07 | [1.02; 1.12] | 0.004 |
| LDL Cholesterol* | Incident | 1.28 | [1.26; 1.30] | <0.0001 |
| | Recurrent | 1.22 | [1.17; 1.28] | <0.0001 |
| HDL Cholesterol* | Incident | 0.72 | [0.70; 0.74] | <0.0001 |
| | Recurrent | 0.99 | [0.94; 1.04] | 0.598 |
| Triglycerides* | Incident | 1.31 | [1.28; 1.33] | <0.0001 |
| | Recurrent | 1.07 | [1.02; 1.12] | 0.003 |
| Body Mass Index | Incident | 1.26 | [1.24; 1.29] | <0.0001 |
| | Recurrent | 1.04 | [1.00; 1.09] | 0.053 |

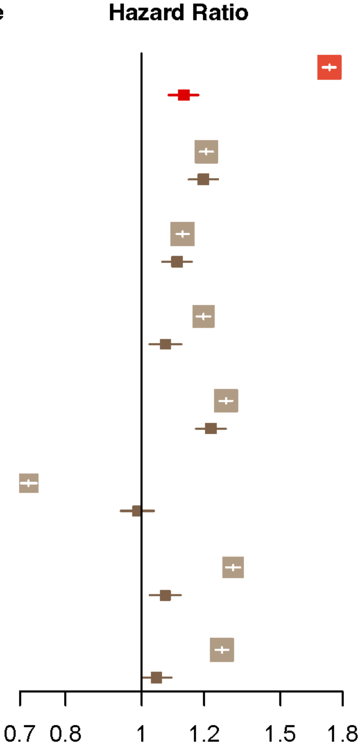

Hazards ratio per standard deviation (HR/SD) of variable of interest for incident disease assessed in individuals without prior coronary artery disease (CAD) followed for development of first CAD event with Cox proportional hazards regression model adjusted for age, sex, genotyping array, and the first ten principal components of ancestry in the European ancestry validation dataset of the UK Biobank Study. Hazards ratio per standard deviation of variable of interest for recurrent disease assessed in individuals with prior CAD followed for development of recurrent CAD event with Cox proportional hazards regression model adjusted for age, sex, genotyping array, and the first ten principal components of ancestry. *LDL-C, HDL-C, and triglyceride values were adjusted for cholesterol-lowering medication status, as previously described.[17] BP: Blood pressure. BMI: Body-mass index. HgbA1c: Glycated hemoglobin. LDL-C: Low-density lipoprotein cholesterol. HDL-C: High-density lipoprotein cholesterol.
